# Accuracy of four lateral flow immunoassays for anti SARS-CoV-2 antibodies: a head-to-head comparative study

**DOI:** 10.1101/2021.01.30.21250777

**Authors:** Hayley E Jones, Ranya Mulchandani, Sian Taylor-Phillips, A E Ades, Justin Shute, Keith R Perry, Nastassya L Chandra, Tim Brooks, Andre Charlett, Matthew Hickman, Isabel Oliver, Stephen Kaptoge, John Danesh, Emanuele Di Angelantonio, COMPARE study investigators, EDSAB-HOME investigators, David Wyllie

## Abstract

**Background:** SARS-CoV-2 antibody tests are used for population surveillance and might have a future role in individual risk assessment. Lateral flow immunoassays (LFIAs) can deliver results rapidly and at scale, but have widely varying accuracy.

**Methods:** In a laboratory setting, we performed head-to-head comparisons of four LFIAs: the Rapid Test Consortium’s AbC-19^™^ Rapid Test, OrientGene COVID IgG/IgM Rapid Test Cassette, SureScreen COVID-19 Rapid Test Cassette, and Biomerica COVID-19 IgG/IgM Rapid Test. We analysed blood samples from 2,847 key workers and 1,995 pre-pandemic blood donors with all four devices.

**Findings:** We observed a clear trade-off between sensitivity and specificity: the IgG band of the SureScreen device and the AbC-19^™^ device had higher specificities but OrientGene and Biomerica higher sensitivities. Based on analysis of pre-pandemic samples, SureScreen IgG band had the highest specificity (98.9%, 95% confidence interval 98.3 to 99.3%), which translated to the highest positive predictive value across any pre-test probability: for example, 95.1% (95%CI 92.6, 96.8%) at 20% pre-test probability. All four devices showed higher sensitivity at higher antibody concentrations (“spectrum effects”), but the extent of this varied by device.

**Interpretation:** The estimates of sensitivity and specificity can be used to adjust for test error rates when using these devices to estimate the prevalence of antibody. If tests were used to determine whether an individual has SARS-CoV-2 antibodies, in an example scenario in which 20% of individuals have antibodies we estimate around 5% of positive results on the most specific device would be false positives.

**Funding:** Public Health England.

**Research in context:** *Evidence before this study:* We searched for evidence on the accuracy of the four devices compared in this study: OrientGene COVID IgG/IgM Rapid Test Cassette, SureScreen COVID-19™ Rapid Test Cassette, Biomerica COVID-19 IgG/IgM Rapid Test and the UK Rapid Test Consortium’s AbC-19™ Rapid Test. We searched Ovid MEDLINE (In-Process & Other Non-Indexed Citations and Daily), PubMed, MedRxiv/BioRxiv and Google Scholar from January 2020 to 16^th^ January 2021. Search terms included device names AND ((SARS-CoV-2) OR (covid)). Of 303 records assessed, data were extracted from 24 studies: 18 reporting on the accuracy of the OrientGene device, 7 SureScreen, 2 AbC-19™ and 1 Biomerica. Only three studies compared the accuracy of two or more of the four devices. With the exception of our previous report on the accuracy of the AbC-19™ device, which the current manuscript builds upon, sample size ranged from 7 to 684. For details, see Supplementary Materials. The largest study compared OrientGene, SureScreen and Biomerica. SureScreen was estimated to have the highest specificity (99.8%, 95% CI 98.9 to 100%) and OrientGene the highest sensitivity (92.6%), but with uncertainty about the latter result due to small sample sizes. The other two comparative studies were small (n = 65, n = 67) and therefore provide very uncertain results. We previously observed spectrum effects for the AbC-19™ device, such that sensitivity is upwardly biased if estimated only from PCR-confirmed cases. The vast majority of previous studies estimated sensitivity in this way.

*Added value of this study:* We performed a large scale (n = 4,842), head-to-head laboratory-based evaluation and comparison of four lateral flow devices, which were selected for evaluation by the UK Department of Health and Social Care’s New Tests Advisory Group, on the basis of a survey of test and performance data available. We evaluated the performance of diagnosis based on both IgG and IgM bands, and the IgG band alone. We found a clear trade-off between sensitivity and specificity across devices, with the SureScreen and AbC-19™ devices being more specific and OrientGene and Biomerica more sensitive. Based on analysis of 1,995 pre-pandemic blood samples, we are 99% confident that SureScreen (IgG band reading) has the highest specificity of the four devices (98.9%, 95% CI 98.3, 99.3%). We found evidence that all four devices have reduced sensitivity at lower antibody indices, i.e. spectrum effects. However, the extent of this varies by device and appears to be less for other devices than for AbC-19. Our estimates of sensitivity and specificity are likely to be higher than would be observed in real use of these devices, as they were based on majority readings of three trained laboratory personnel.

*Implications of all the available evidence:* When used in epidemiological studies of antibody prevalence, the estimates of sensitivity and specificity provided in this study can be used to adjust for test errors. Increased precision in error rates will translate to increased precision in seroprevalence estimates. If lateral flow devices were used for individual risk assessment, devices with maximum specificity would be preferable. However, if, for an example, 20% of the tested population had antibodies, we estimate that around 1 in 20 positive results on the most specific device would be incorrect.

## Introduction

Tests for SARS-CoV-2 antibodies are used for population serosurveillance (1, 2) and could in future be used for post-vaccination seroepidemiology. With emerging evidence of antibodies being associated with reduced risk of COVID-19 disease (3-7), antibody tests might also have a role in individual risk assessment (8), pending improved understanding of the mechanisms and longevity of immunity. Both uses require understanding of test sensitivity and specificity: these can be used to adjust seroprevalence estimates for test errors (9), while any test used for individual risk assessment would need to be shown to be sufficiently accurate, in particular, highly specific (10, 11).

A number of laboratory-based immunoassays and lateral flow immunoassays (LFIAs) are now available, which detect IgG and/or IgM responses to the spike or nucleoprotein antigens (12-14). Following infection with SARS-CoV-2, most individuals generate antibodies against both of these antigens (15). Existing efficacious recombinant vaccines contain the spike antigen (16), therefore vaccinated individuals generate only a response to this. LFIAs are small devices which produce results rapidly, without the need for a laboratory, and therefore have the potential to be employed at scale.

A Cochrane review found 38 studies evaluating LFIAs for SARS-CoV-2 antibodies already by late April 2020. However, results from most studies were judged to be at high risk of bias, and very few studies directly compared multiple devices (14). Where direct comparisons have been performed, they have shown that accuracy of LFIAs varies widely across devices (13, 17-20). A key limitation of most studies is that sensitivity has been estimated only from individuals who previously had a positive PCR test. In a recent evaluation of one LFIA, the UK Rapid Test Consortium’s “AbC-19™ Rapid Test” (21) (AbC-19 hereafter), we found evidence that this can over-estimate sensitivity (21). We attributed this to PCR-confirmed cases tending to be more severe – particularly early in the pandemic, when testing access was very limited – and producing a greater antibody response, i.e. “spectrum bias” (22, 23).

In this paper, we present a head-to-head comparison of the accuracy of AbC-19, and three other LFIAs, based on a large (n = 4,842) number of blood samples. The three additional devices were OrientGene “COVID IgG/IgM Rapid Test Cassette”, SureScreen “COVID-19 Rapid Test Cassette”, and Biomerica “COVID-19 IgG/IgM Rapid Test”, hereafter referred to as OrientGene, SureScreen and Biomerica for brevity.

## Methods

We analysed blood samples from 2,847 key workers participating in the EDSAB-HOME study and 1,995 pre-pandemic blood donors from the COMPARE study (24), in a laboratory setting. All samples were from distinct individuals. We evaluated each device using two approaches. First (Approach 1), we compared LFIA results with the known previous infection status of pre-pandemic blood donors (“known negatives”) and 268 EDSAB-HOME participants reporting previous PCR positivity (“known positives”). Second (Approach 2), we compared LFIA results with results on two sensitive laboratory immunoassays in EDSAB-HOME participants. We have previously reported accuracy of the AbC-19 device based on the same sample set and overall approaches: these results are reproduced here for comparative purposes (21).

### Lateral flow immunoassays

Devices (Table S1) were selected by the UK Department of Health and Social Care’s New Tests Advisory Group, on the basis of a survey of test and performance data available. AbC-19, OrientGene and SureScreen devices contain SARS-CoV-2 Spike protein, or domains from it, while Biomerica contains Nucleoprotein. All four devices give qualitative positive or negative results. AbC-19 detects IgG only, while the other three devices contain separate bands representing detection of IgG and IgM. We report results for these by two different scoring strategies: (i) “one band”, in which we considered a result to be positive only if the IgG band was positive; and (ii) “two band”, in which we considered results to be positive if either band was positive. In statistical analysis, these two readings were treated as separate “tests”, such that our comparison was of seven tests in total. By definition, the “two band” reading of each device has sensitivity greater than or equal to, but specificity less than or equal to, the “one band” reading.

### Study participants

Study participants (n = 4,842), including baseline characteristics, are described in full elsewhere (21). A flow diagram is provided (Figure S1).

EDSAB-HOME (ISRCTN56609224) participants were key workers (healthcare workers, fire and police officers) in England, who had a venous blood sample taken at a study clinic in June 2020. Individuals in recruitment Streams A and B (n = 2,693) were recruited without regard to previous SARS-CoV-2 infection status, while Stream C (n = 154) participants were recruited based on self-reported previous PCR positivity. Some Stream A/B individuals (n = 114) also self-reported previous PCR positivity. We refer to the total (n = 268) individuals who self-reported a previous PCR positive result as “known positives” and to the remaining n = 2,579 EDSAB-HOME participants as “individuals with unknown previous infection status” at clinic visit. See Supplementary Materials for more information.

COMPARE (ISRCTN90871183) was a 2016-2017 blood donor cohort study in England (24). We performed stratified random sampling, by age, sex and region to select 2,000 participants, of whom 1,995 had samples available for analysis. We refer to these samples as “known negatives”.

### Laboratory protocol

Tests were performed by experienced laboratory staff. All EDSAB-HOME samples were first tested with two laboratory immunoassays: Roche Elecsys®, which measures total (including IgG and IgM) antibodies against the Nucleoprotein, and EuroImmun, which measures IgG antibodies against the S protein S1 domain. We used thresholds of 1.0 for Roche Elecsys® and 0.8 for EuroImmun (manufacturer recommended positive and “borderline” thresholds, respectively).

Each device was independently read by three members of staff. Readers were blind to demographic or clinical information on participants and to results on any previous assays. Each reader scored each test band using the WHO scoring system for subjectively read assays, in which 0 represents “negative”, 1 “very weak but definitely reactive”, 2 “medium to strong reactivity” and 7 “invalid” (25). The majority reading was taken as the final result. For assessment of test sensitivity and specificity, scores of 1 and 2 were grouped as “positive”. If any band of an LFIA device was assessed as “invalid” on initial testing, the sample was re-tested. In this situation, we treat the re-test result as primary. We also report, but as secondary, results following re-testing of apparent errors: see Supplementary Materials.

### Estimation of sensitivity, specificity, positive and negative predictive values

Approach 1: We estimated LFIA accuracy through comparison of results with the known previous SARS-CoV-2 infection status of individuals. Specificity was estimated from all 1,995 “known negative” samples. The association of false positivity with age, sex and ethnicity was also explored. We estimated sensitivity from the 268 “known positive” EDSAB-HOME samples. Numbers of false negatives are also reported by time since symptom onset and separately for asymptomatic individuals.

Approach 2: We then estimated LFIA accuracy through comparison with results on the Roche Elecsys® laboratory immunoassay in EDSAB-HOME samples. This assay has been estimated to have sensitivity of 97.2% (95% CI 95.4, 98.4%) and specificity of 99.8% (99.3, 100%) to previous infection (12). As sensitivity analyses, we also report accuracy estimates based on comparison with EuroImmun and a composite reference standard of “positive on either laboratory assay versus negative on both”. Estimates of sensitivity were calculated separately for known positives and individuals with unknown previous infection status, to assess for potential spectrum bias (21). Specificity was estimated from reference standard negative individuals among the “unknown previous infection status” population. As EDSAB-HOME Streams A and B comprise a “one gate” population (21, 26), we also report Approach 2 results from all EDSAB-HOME streams A and B participants combined, regardless of previous PCR positivity.

Positive and negative predictive values: We estimated the positive and negative predictive value (PPV and NPV) for example scenarios of a 10%, 20% and 30% pre-test probability. To calculate these, we used estimates of specificity based on pre-pandemic sera (Approach 1) and sensitivity based on comparison with Roche Elecsys® in individuals with unknown previous infection status (Approach 2). These choices were based on an expectation (following our previous work, 21) that these estimates of the respective parameters would be the least susceptible to bias.

### Statistical analysis

Statistical analysis was performed in R4.0.3 and Stata 15. Sensitivity and specificity were estimated by observed proportions based on each reference standard, with 95% CIs computed using Wilson’s method. Logistic regressions with age, sex and ethnicity as covariates were used to explore potential associations with false positivity. To further explore potential associations with age, we also fitted fractional polynomials and plotted the best fitting functional form for each test.

In comparing the sensitivity and specificity of the seven “tests”, we used generalised estimating equations (GEE) to account for conditional correlations among results (27). For example, in Approach 1 we fitted separate GEE regressions, with test as a covariate, to the “known positives” and to the “known negatives”. We used independence working covariance matrices (27).

We obtained 95% CIs around PPVs, NPVs, differences in sensitivity and differences in specificity using Monte Carlo simulation. We sampled from a multivariate normal distribution for GEE regression coefficients, using the parameter estimates and robust variance-covariance matrix. We also present ranks (from 1 to 7) for each set of sensitivity, specificity and PPV estimates. Ranks were calculated at each iteration of the Monte Carlo simulations, and summarised by medians and 95% CIs across simulations (28). We further report the proportion of simulations for which each test was ranked first.

### Assessment of spectrum effects in test sensitivity

Within the Approach 2 analysis of Roche Elecsys® positives, we report the absolute difference between sensitivity estimated from PCR-confirmed cases and sensitivity estimated from individuals with unknown previous infection status, with 95% CI. Among individuals who were positive on Roche Elecsys®, we also plotted the relationship between sensitivity and anti-Nucleoprotein (Roche Elecsys®) and anti-S1 (EuroImmun) antibody indices. To aid visual assessment of these relationships, we fitted dose-response relationships (using R drc package), with model selection based on the Akaike information criterion.

### Role of the funding source

The study was commissioned by the UK Government’s Department of Health and Social Care (DHSC), and was funded and implemented by Public Health England, supported by the NIHR Clinical Research Network Portfolio. The DHSC had no role in the study design, data collection, analysis, interpretation of results, writing of the manuscript, or the decision to publish.

## Results

### Main results

Figures 1 and 2 show results from Approach 1 and from the Approach 2 analysis of individuals with unknown previous infection status at clinic visit, which are mutually exclusive and exhaustive subsets of the 4,842 samples. Also shown are results from the Approach 2 sensitivity analyses with alternative reference standards. Estimated differences between the sensitivity and specificity of tests, with 95% CIs, are shown in Tables S2 and S3.

**Figure 1:**
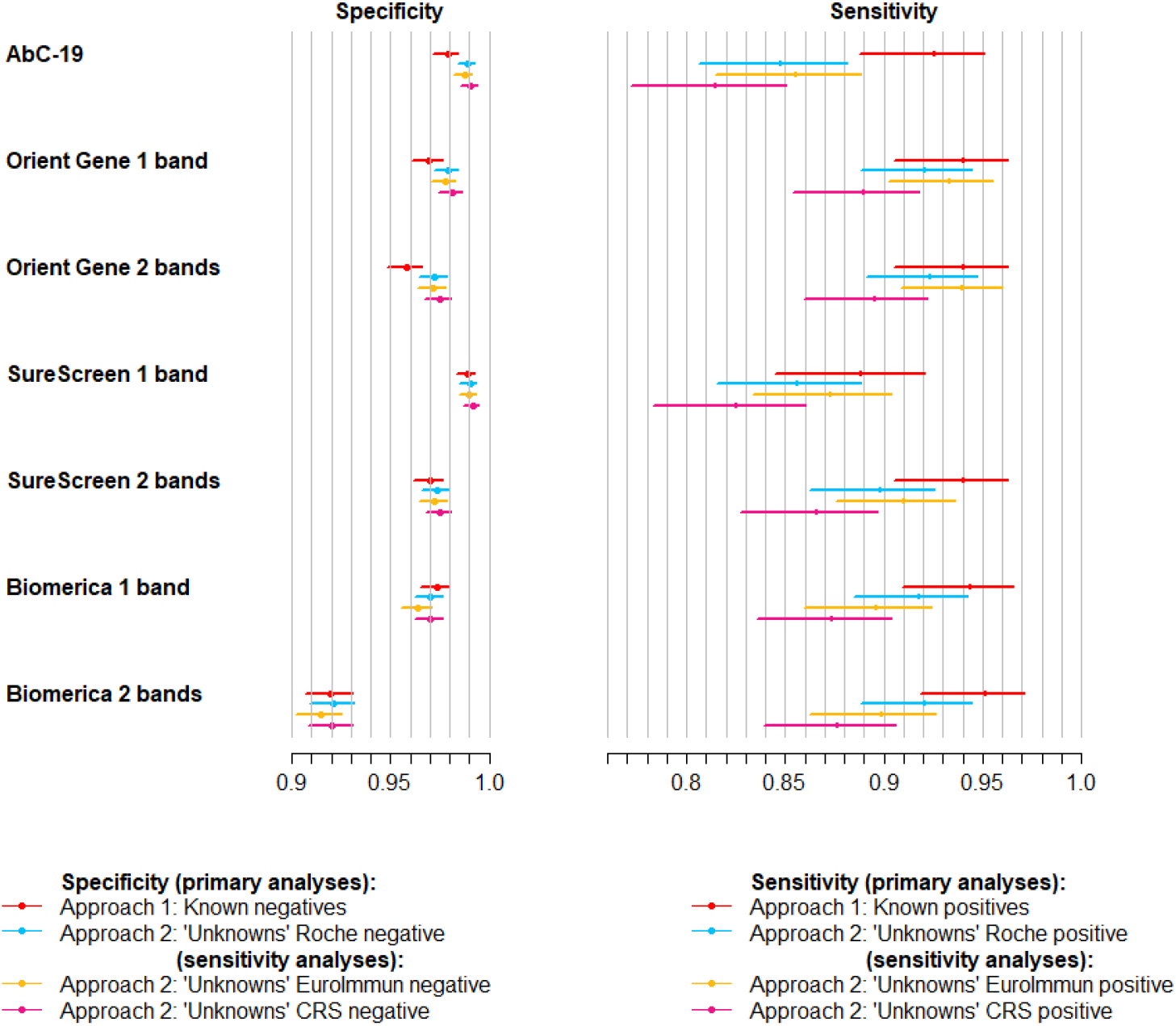
Sensitivity and specificity of lateral flow devices, with 95% confidence intervals. Four sets of estimates are shown: (i) Approach 1, i.e. specificity from analysis of 1,995 known negatives and sensitivity from 268 known positives; (ii) Approach 2 analysis of 2,579 individuals with unknown previous infection status (“unknowns”), calculated against Roche Elecsys reference standard; (iii) Approach 2 sensitivity analysis: analysis of unknowns compared with alternative EuroImmun reference standard; (iv) Approach 2 sensitivity analysis: analysis of unknowns compared with alternative composite reference standard (CRS) of positive on either Roche Elecsys or EuroImmun versus negative on both.

**Figure 2:**
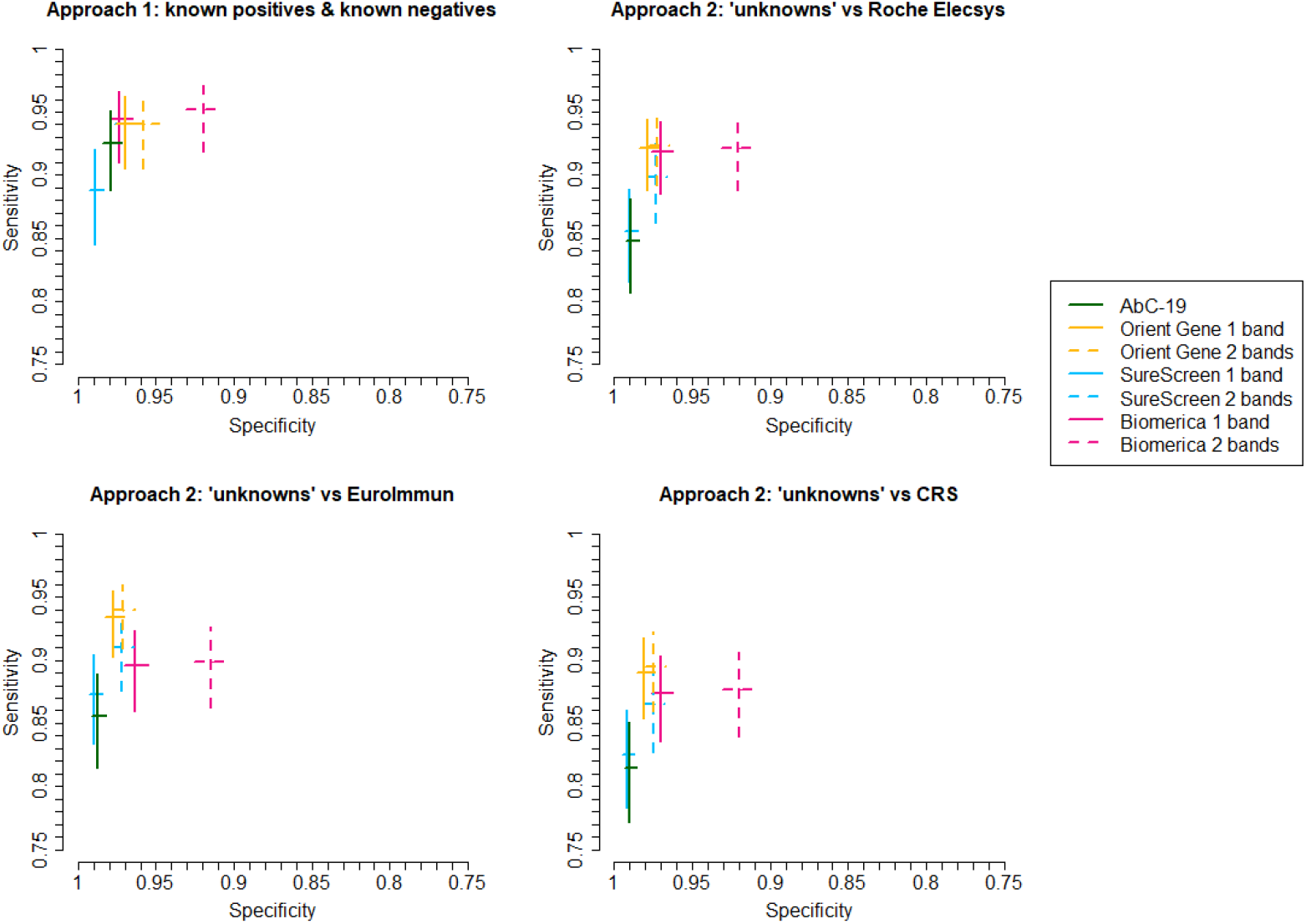
Sensitivity and specificity of lateral flow devices, with 95% confidence intervals, plotted in Receiver Operator Characteristic space. Four sets of estimates are shown: (i) Approach 1, i.e. specificity from analysis of 1,995 known negatives and sensitivity from 268 known positives; (ii) Approach 2 analysis of 2,579 individuals with unknown previous infection status (“unknowns”), calculated against Roche Elecsys reference standard; (iii) Approach 2 sensitivity analysis: analysis of unknowns compared with alternative EuroImmun reference standard; (iv) Approach 2 sensitivity analysis: analysis of unknowns compared with alternative composite reference standard (CRS) of positive on either Roche Elecsys or EuroImmun versus negative on both. NB SureScreen 2 band overlays Orient Gene 1 band in the first panel.

Both approaches show a clear trade-off between sensitivity and specificity, with SureScreen 1 band and AbC-19 having higher specificities but lowest sensitivities, while OrientGene and Biomerica have higher sensitivities but lower specificities.

From Approach 1, SureScreen 1 band is estimated to have higher specificity but lower sensitivity than AbC-19, whereas the two tests appeared comparable (although with all point estimates marginally favouring SureScreen) from Approach 2. Resulting from this, we estimate the one band reading of the SureScreen device to have the highest PPV.

### Approach 1: comparison with known previous infection status

Approach 1 estimates are shown in Tables 1 (specificity) and 2 (sensitivity).

**Table 1:** Specificity of lateral flow devices: Approach 1 (known negatives). Estimates based on analysis of 1,995 pre-pandemic samples. CI = confidence interval, TNs = true negatives, FPs = false positives, “Probability best” = the proportion of Monte Carlo simulations in which the test had the highest specificity. Note: these AbC-19 ™ results have been published previously (21) and are reproduced here for comparative purposes.

|  | Lateral flow immunoassay |  |  |  |  |  |  |
| --- | --- | --- | --- | --- | --- | --- | --- |
|  | AbC-19™ | Orient Gene 1 band | Orient Gene 2 bands | SureScreen 1 band | SureScreen 2 bands | Biomerica 1 band | Biomerica 2 bands |
| TNs | 1,953 | 1,934 | 1,911 | 1,973 | 1,935 | 1,942 | 1,835 |
| FPs | 42 | 61 | 84 | 22 | 60 | 53 | 160 |
| Specificity<br>(95% CI) | 97.9%<br>(97.2, 98.4) | 96.9%<br>(96.1, 97.6) | 95.8%<br>(94.8, 96.6) | 98.9%<br>(98.3, 99.3) | 97.0%<br>(96.1, 97.7) | 97.3%<br>(96.5, 98.0) | 92.0%<br>(90.7, 93.1) |
| Rank<br>(95% CI) | 2<br>(2, 4) | 4<br>(3, 5) | 6<br>(6, 6) | 1<br>(1, 1) | 4<br>(3, 5) | 3<br>(2, 5) | 7<br>(7, 7) |
| Probability best | 0.01 | 0.00 | 0.00 | 0.99 | 0.00 | 0.00 | 0.00 |

SureScreen 1 band reading was estimated to have 98.9% specificity (95% CI 98.3, 99.3%), with high certainty (99%) of this being the highest. This is 1.0% (95% CI 0.2, 1.8%) higher than the specificity of AbC-19 (Table S2), which was ranked 2^nd^ (95% CI 2 ^nd^, 4^th^). There was no strong evidence of any association between false positivity and age for any device (Table S3) although there was some indication that Biomerica 1 band specificity might decline in older adults (Figure S2). With the exception of an apparent association of false positivity of the AbC-19 device with sex, which we have reported previously (21), there was no indication of specificity varying by sex or ethnicity (Table S4). SureScreen 1 band was, however, estimated to have the lowest sensitivity when this was estimated from PCR-confirmed cases only (Table 2: 88.8%, 95% CI 84.5, 92.0%), 3.7% (95% CI 0.5, 7.1%) lower than AbC-19 (Table S5).

**Table 2:** Sensitivity of lateral flow devices: Approach 1 (known positives). Estimates based on analysis of 268 individuals self-reporting previous PCR confirmed infection. CI = confidence interval, TPs = true positives, FNs = false negatives, “Probability best” = the proportion of Monte Carlo simulations in which the test had the highest sensitivity. Note: these AbC-19 ™ results have been published previously (21) and are reproduced here for comparative purposes.

|  | Lateral flow immunoassay |  |  |  |  |  |  |
| --- | --- | --- | --- | --- | --- | --- | --- |
|  | AbC-19™ | Orient Gene 1 band | Orient Gene 2 bands | SureScreen 1 band | SureScreen 2 bands | Biomerica 1 band | Biomerica 2 bands |
| TPs | 248 | 252 | 252 | 238 | 252 | 253 | 255 |
| FNs | 20 | 16 | 16 | 30 | 16 | 15 | 13 |
| Sensitivity (95% CI) | 92.5% (88.8, 95.1) | 94.0% (90.5, 96.3) | 94.0% (90.5, 96.3) | 88.8% (84.5, 92.0) | 94.0% (90.5, 96.3) | 94.4% (91.0, 96.6) | 95.1% (91.9, 97.1) |
| Rank (95% CI) | 6 (2, 6) | 4 (1, 6) | 4 (1, 6) | 7 (7, 7) | 3 (1, 6) | 3 (1, 6) | 1 (1, 5) |
| Probability best | 0.01 | 0.08 | 0.08 | 0.00 | 0.14 | 0.04 | 0.64 |
| *False negatives by days since symptom onset to test:* |  |  |  |  |  |  |  |
| Asymptomatic (n=12) | 5 | 3 | 3 | 5 | 4 | 4 | 4 |
| 8-21 days (n=5) | 0 | 0 | 0 | 0 | 0 | 0 | 0 |
| 22-35 days (n=20) | 1 | 1 | 1 | 2 | 1 | 1 | 1 |
| 36-70 days (n=142) | 6 | 4 | 4 | 12 | 5 | 2 | 1 |
| ≥71 days (n=89) | 8 | 8 | 8 | 11 | 6 | 8 | 7 |

### Approach 2: comparison with laboratory immunoassay results in EDSAB-HOME samples

Among the 268 “known positives”, nine were negative on Roche Elecsys®. Removing these from the denominator slightly increased point estimates of sensitivity (Table 3), but had no notable impact on rankings. Among the 2,579 individuals with unknown previous infection status, 354 were positive on Roche Elecsys®. Point estimates of sensitivity were lower for all seven tests in this population than among known positives (see below). In this population, there was evidence that both the OrientGene and Biomerica devices have higher sensitivity than SureScreen or AbC-19 (Table S5). There was no evidence of a difference between the sensitivity of SureScreen and AbC-19 (absolute difference in favour of SureScreen = 0.8%, 95% CI −2.2, 3.9%). Increases in sensitivity in the 2 band versus 1 band reading of OrientGene and Biomerica devices were minimal.

**Table 3:** Sensitivity and specificity of lateral flow devices: Approach 2. Comparison with Roche Elecsys immunoassay in EDSAB-HOME samples, stratified by previous PCR positivity. CI = confidence interval, “Probability best” = the proportion of Monte Carlo simulations in which the test had the highest sensitivity or specificity. Note: the AbC-19 ™ results have been published previously (21) and are reproduced here for comparative purposes.

|  | AbC-19™ | Orient Gene 1<br>band | Orient Gene 2<br>bands | SureScreen 1<br>band | SureScreen 2<br>bands | Biomerica 1<br>band | Biomerica 2<br>bands |
| --- | --- | --- | --- | --- | --- | --- | --- |
| **Analysis of 268 PCR-confirmed cases** |  |  |  |  |  |  |  |
| Reference standard of Roche Elecsys: 259 positive |  |  |  |  |  |  |  |
| False negatives | 15 | 12 | 12 | 23 | 10 | 8 | 6 |
| Sensitivity (95% CI) | 94.2%<br>(90.7, 96.5) | 95.4%<br>(92.1, 97.3) | 95.4%<br>(92.1, 97.3) | 91.1%<br>(87.0, 94.0) | 96.1%<br>(93.0, 97.9) | 96.9%<br>(94.0, 98.4) | 97.7%<br>(95.0, 98.9) |
| Ranked sensitivity (95% CI) | 6 (3, 7) | 4 (2, 6) | 4 (2, 6) | 7 (6, 7) | 3 (1, 6) | 2 (1, 6) | 1 (1, 5) |
| Probability best sensitivity | 0.00 | 0.01 | 0.01 | 0.00 | 0.14 | 0.05 | 0.79 |
| **Analysis of 2,579 individuals with unknown previous infection status** |  |  |  |  |  |  |  |
| Reference standard of Roche Elecsys: 354 positive and 2,225 negative |  |  |  |  |  |  |  |
| False negatives | 54 | 28 | 27 | 51 | 36 | 29 | 28 |
| Sensitivity (95% CI) | 84.7%<br>(80.6, 88.1) | 92.1%<br>(88.8, 94.5) | 92.4%<br>(89.1, 94.7) | 85.6%<br>(81.6, 88.9) | 89.8%<br>(86.2, 92.6) | 91.8%<br>(88.5, 94.2) | 92.1%<br>(88.8, 94.5) |
| Ranked sensitivity (95% CI) | 7 (6, 7) | 2 (1, 4) | 1 (1, 4) | 6 (6, 7) | 5 (3, 5) | 4 (1, 5) | 3 (1, 4) |
| Probability best sensitivity | 0.00 | 0.07 | 0.50 | 0.00 | 0.00 | 0.06 | 0.36 |
| False positives | 24 | 47 | 62 | 22 | 59 | 66 | 176 |
| Specificity (95% CI) | 98.9%<br>(98.4, 99.3) | 97.9%<br>(97.2, 98.4) | 97.2%<br>(96.4, 97.8) | 99.0%<br>(98.5, 99.3) | 97.3%<br>(96.6, 97.9) | 97.0%<br>(96.2, 97.7) | 92.1%<br>(90.9, 93.1) |
| Ranked specificity (95% CI) | 2 (1, 2) | 3 (3, 4) | 5 (4, 6) | 1 (1, 2) | 4 (3, 6) | 6 (3, 6) | 7 (7, 7) |
| Probability best specificity | 0.37 | 0.00 | 0.00 | 0.63 | 0.00 | 0.00 | 0.00 |
| Absolute difference in sensitivity: known positives vs individuals with unknown previous infection status (95% CI) | 9.5%<br>(7.7, 11.3) | 3.3%<br>(-0.2, 6.6) | 3.0%<br>(-0.5, 6.3) | 5.5%<br>(2.2, 8.7) | 6.3%<br>(3.0, 9.7) | 5.1%<br>(2.3, 8.0) | 5.6%<br>(3.0, 8.3) |

Based on the 2,225 individuals with unknown previous infection status who were negative on Roche Elecsys®, specificity estimates were very similar to those from Approach 1 for SureScreen and Biomerica, but around 1% higher for AbC-19 and OrientGene (Table 3). The ranking of devices was consistent across the two approaches, but the observed difference in specificity between SureScreen and AbC-19 was much reduced in Approach 2, with a wide CI (difference = 0.1%, 95% CI −0.4 to 0.6%, Table S2).

Figures 1, 2 and Tables S6, S7 show results from sensitivity analyses on the 2,579 samples from individuals with unknown previous infection status. Estimates of specificity were robust, while sensitivity appeared slightly higher for AbC-19, OrientGene and SureScreen, but slightly lower for Biomerica, when EuroImmun was taken as the reference standard, although with overlapping CIs. All devices were estimated to have slightly lower sensitivity when evaluated against the CRS. OrientGene was ranked highest for sensitivity across all three immunoassay reference standards, but with Biomerica appearing as a close contender when evaluated against Roche Elecsys®. Table S8 shows sensitivity and specificity estimated from all EDSAB-HOME Streams A and B (“one gate” study), based on comparison with each of the three immunoassay reference standards. Rankings of devices were quite robust to inclusion of PCR-confirmed cases.

Re-test results are shown in Table S9.

### Positive and negative predictive values

Based on the sets of estimates that we consider least susceptible to bias (see Methods), we are 99% confident that SureScreen 1 band reading has the highest PPV. This ranking does not depend on pre-test probability (Table 4, Figure S4). At a pre-test probability of 20%, we estimate SureScreen 1 band reading to have a PPV of 95.1% (92.6, 96.8%), such that we would expect approximately one in twenty positive results to be incorrect.

**Table 4:**
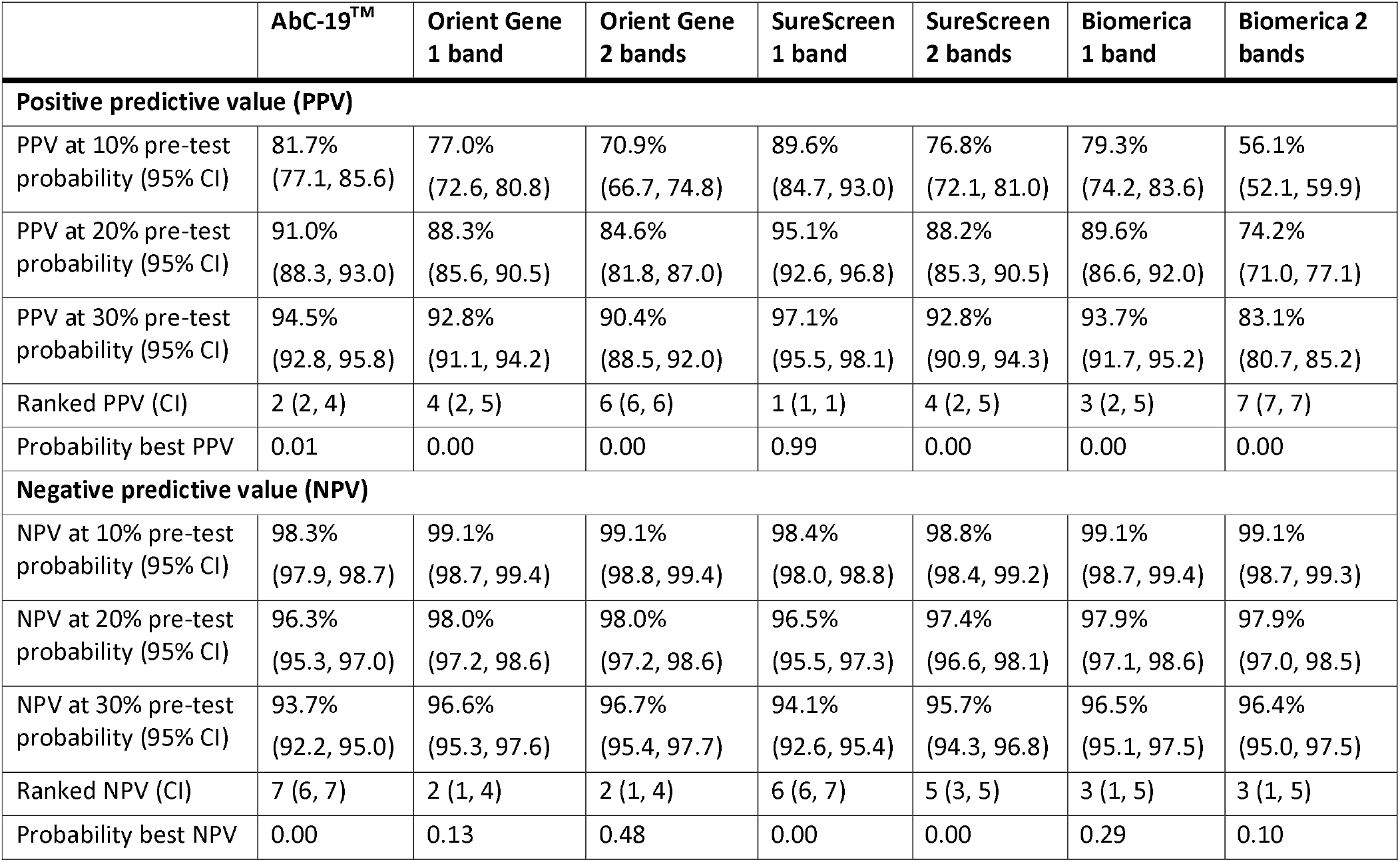
Positive predictive value (PPV) and negative predictive value (NPV), with 95% confidence intervals (CIs), for example scenarios of 10%, 20% and 30% pre-test probability. Probability best” = the proportion of Monte Carlo simulations in which the test had the highest PPV or NPV. Specificity estimated from 1,995 pre-pandemic samples (Table 1) and sensitivity from 354 Roche Elecsys positives with unknown previous infection status at clinic visit (Table 3).

|  | **AbC-19™** | **Orient Gene**<br>**1 band** | **Orient Gene**<br>**2 bands** | **SureScreen**<br>**1 band** | **SureScreen**<br>**2 bands** | **Biomerica**<br>**1 band** | **Biomerica 2**<br>**bands** |
| --- | --- | --- | --- | --- | --- | --- | --- |
| **Positive predictive value (PPV)** |  |  |  |  |  |  |  |
| PPV at 10% pre-test probability (95% CI) | 81.7%<br>(77.1, 85.6) | 77.0%<br>(72.6, 80.8) | 70.9%<br>(66.7, 74.8) | 89.6%<br>(84.7, 93.0) | 76.8%<br>(72.1, 81.0) | 79.3%<br>(74.2, 83.6) | 56.1%<br>(52.1, 59.9) |
| PPV at 20% pre-test probability (95% CI) | 91.0%<br>(88.3, 93.0) | 88.3%<br>(85.6, 90.5) | 84.6%<br>(81.8, 87.0) | 95.1%<br>(92.6, 96.8) | 88.2%<br>(85.3, 90.5) | 89.6%<br>(86.6, 92.0) | 74.2%<br>(71.0, 77.1) |
| PPV at 30% pre-test probability (95% CI) | 94.5%<br>(92.8, 95.8) | 92.8%<br>(91.1, 94.2) | 90.4%<br>(88.5, 92.0) | 97.1%<br>(95.5, 98.1) | 92.8%<br>(90.9, 94.3) | 93.7%<br>(91.7, 95.2) | 83.1%<br>(80.7, 85.2) |
| Ranked PPV (CI) | 2 (2, 4) | 4 (2, 5) | 6 (6, 6) | 1 (1, 1) | 4 (2, 5) | 3 (2, 5) | 7 (7, 7) |
| Probability best PPV | 0.01 | 0.00 | 0.00 | 0.99 | 0.00 | 0.00 | 0.00 |
| **Negative predictive value (NPV)** |  |  |  |  |  |  |  |
| NPV at 10% pre-test probability (95% CI) | 98.3%<br>(97.9, 98.7) | 99.1%<br>(98.7, 99.4) | 99.1%<br>(98.8, 99.4) | 98.4%<br>(98.0, 98.8) | 98.8%<br>(98.4, 99.2) | 99.1%<br>(98.7, 99.4) | 99.1%<br>(98.7, 99.3) |
| NPV at 20% pre-test probability (95% CI) | 96.3%<br>(95.3, 97.0) | 98.0%<br>(97.2, 98.6) | 98.0%<br>(97.2, 98.6) | 96.5%<br>(95.5, 97.3) | 97.4%<br>(96.6, 98.1) | 97.9%<br>(97.1, 98.6) | 97.9%<br>(97.0, 98.5) |
| NPV at 30% pre-test probability (95% CI) | 93.7%<br>(92.2, 95.0) | 96.6%<br>(95.3, 97.6) | 96.7%<br>(95.4, 97.7) | 94.1%<br>(92.6, 95.4) | 95.7%<br>(94.3, 96.8) | 96.5%<br>(95.1, 97.5) | 96.4%<br>(95.0, 97.5) |
| Ranked NPV (CI) | 7 (6, 7) | 2 (1, 4) | 2 (1, 4) | 6 (6, 7) | 5 (3, 5) | 3 (1, 5) | 3 (1, 5) |
| Probability best NPV | 0.00 | 0.13 | 0.48 | 0.00 | 0.00 | 0.29 | 0.10 |

OrientGene and Biomerica have the highest ranking NPVs. There is very little difference between the NPVs for the one or two band readings of these devices (Table 4).

### Spectrum effects

For all seven tests, point estimates for sensitivity were lower among individuals with unknown previous infection status who were positive on Roche Elecsys® than among PCR-confirmed cases, with strong statistical evidence of a difference for all tests except OrientGene (Table 3). The greatest observed difference was for AbC-19.

Figure 3 shows that all devices were more sensitive at higher antibody concentrations. This effect was most marked in the devices with lower sensitivity, particularly AbC-19. All LFIAs had high sensitivity at the highest anti-S IgG concentrations, but at lower concentrations many lateral flow tests were falsely negative (Figure S3).

**Figure 3:**
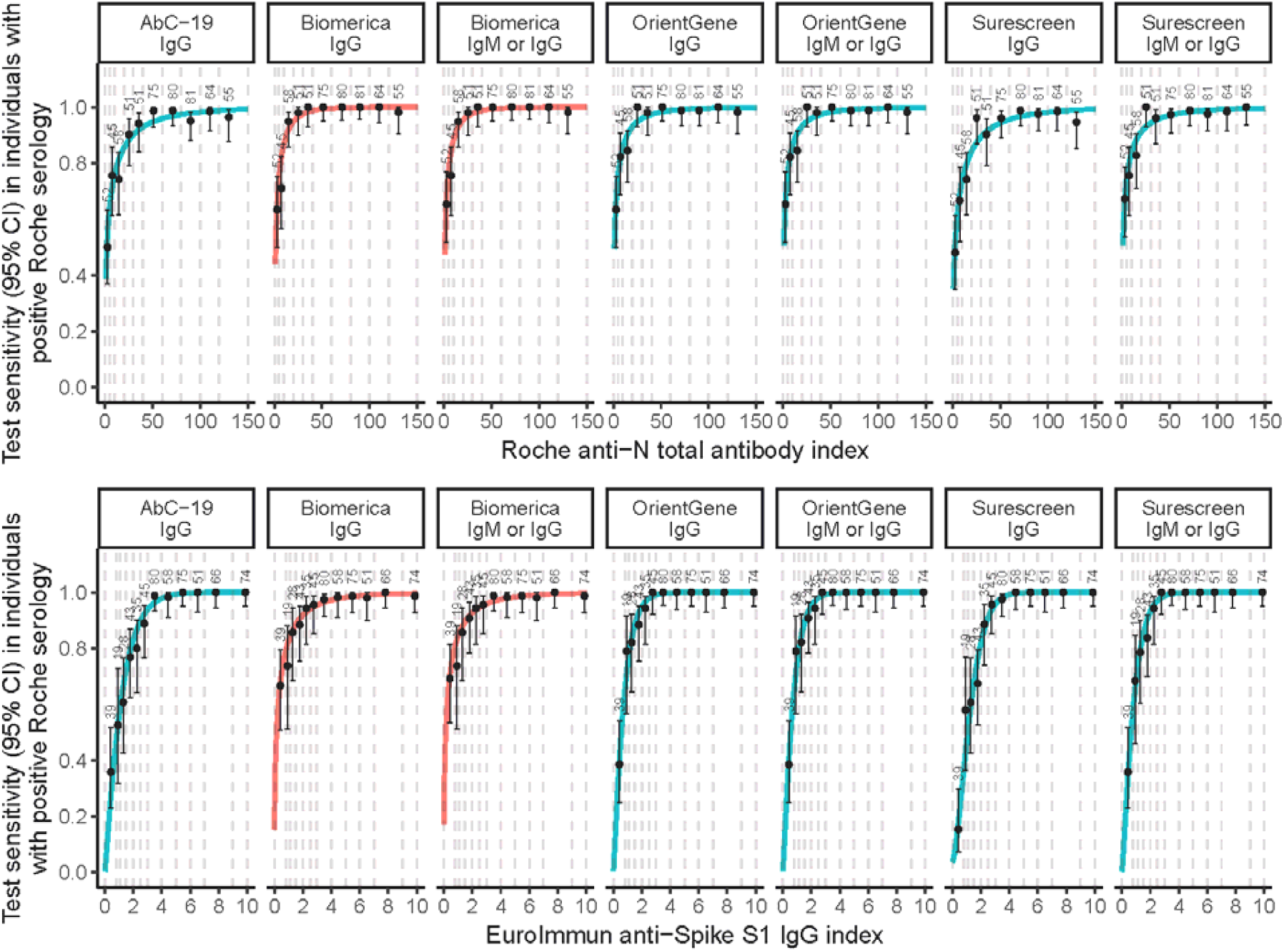
Sensitivity of lateral flow devices, with 95% confidence intervals, by antibody index (categorised into groups of approximately equal size), among n = 613 EDSAB-HOME participants who were positive on Roche Elecsys. Top panel: sensitivity by anti-Nucleoprotein antibody (Roche Elecsys); Bottom Panel: sensitivity by anti-Spike IgG (EuroImmun). Red lines: test contains nucleoprotein; Blue lines: test contains spike proteins.

### Usability issues

Very few bands or devices produced invalid readings (Table S10). Laboratory assessors reported that SureScreen bands were intense, well defined and easy to read, and that OrientGene bands were also easy to read. For Biomerica, some gradients and streaking in band formation was noticed, which led to devices taking slightly longer to read. As we have reported previously, AbC-19 bands were often weak visually (21).

All devices showed some variability in reading across three assessors. Concordance was highest for the SureScreen IgG band: there were no discrepancies in the reading of this for 98.7% (98.3, 98.9%) of devices (Table S11). Positive OrientGene, Biomerica and SureScreen IgG bands all tended to be stronger than AbC-19 bands: for example, across all 613 EDSAB-HOME samples that were positive on Roche Elecsys®, 76%, 69% and 79% showed “medium to strong reactivity” respectively, compared with 44% of AbC-19 devices (Table S12). Concordance was lower for reading of IgM than IgG bands. IgM bands, when read as positive, were also often weak.

## Discussion

We found evidence that SureScreen (when reading the IgG band only) and AbC-19 had higher specificities than OrientGene and Biomerica, but the latter have higher sensitivities. We can confidently conclude that SureScreen 1 band reading has ∼99% specificity, since this estimate was robust across two large discrete samples sets. In contrast, estimates of the specificity of AbC-19 and OrientGene varied slightly across Approaches 1 and 2. As Approach 2 denominators are subject to some misclassification error, we consider the estimates of specificity based on pre-pandemic samples to be most reliable.

The sensitivities of OrientGene and Biomerica appeared comparable based on a reference standard of Roche Elecsys® (anti-N) immunoassay, whereas OrientGene appeared to have higher sensitivity when an alternative (anti-S) reference standard was used. This difference is not surprising since Biomerica also measures anti-N response whereas OrientGene (and the other two devices studied) measures anti-S response. For all four devices, there was some evidence of lower sensitivity to detect lower concentrations of antibody. This spectrum effect appeared strongest for the AbC-19 test and weakest for OrientGene. Due to spectrum effects, we consider Approach 2 estimates of sensitivity to be the most realistic.

Notably, none of the four devices met the UK Medicines and Health Products Regulatory Agency’s requirement of sensitivity >98% for the use case of individual level risk assessment (11), even in our least conservative analytical scenario, which we expect to over-estimate sensitivity. On the other hand, the basis for this criterion is unclear, as we would expect high specificity to be the key consideration for this potential use case.

The major strengths of this work are its size and performance of all LFIAs on an identical sample set. This design is optimal for comparing test accuracy (29). Inclusion of laboratory immunoassay positive cases without PCR confirmation allowed assessment of spectrum effects. A limitation is that tests were conducted in a laboratory setting, with the majority reading across three expert readers being taken as the result. For devices with discrepancies between readers, the accuracy of a single reader can be expected to lower (21). Accuracy may be lower still if devices were read by individuals with less or no training, and may differ if device reading technologies were used. SureScreen IgG band, followed by OrientGene IgG band, had the highest concordance across readers.

An ongoing difficulty in this field is the ambiguity as to whether the true parameters of interest are sensitivity and specificity to previous infection, to presence of specific antibodies, or to “immunity”. Although most individuals seroconvert (15), both the anti-S and anti-N antibody response appears to be highly specific to SARS-CoV-2 (12), and there is now evidence that presence of antibody response correlates with reduced risk (3, 4), these three potential “target conditions” are unlikely to coincide exactly, particularly given declining antibody responses over time. Our estimates are best interpreted as sensitivity and specificity to “recent” SARS-CoV-2 infection (Approach 1) or to presence of an antibody response (Approach 2), which we expect to correlate very highly. Estimates of sensitivity based on a laboratory immunoassay reference standard may be slightly biased due to errors made by the reference standard. We explored this with sensitivity analyses using two alternative reference standards.

Our study describes test accuracy following natural infection, not after vaccination. Estimates of sensitivity would require further validation in vaccinated populations, if the tests were to be used for post-vaccination monitoring. Notably, antigen choice precludes both Biomerica and Roche Elecsys from this use case. Further, we did not quantify the accuracy of tests used in sequence, e.g. check positive results on Test A with a confirmatory Test B (30).

If these devices are used for seroprevalence estimation, our estimates of test accuracy can be used to adjust for test errors (9). The “one gate” estimates of sensitivity would likely be the most appropriate for this. For the alternative potential use case of individual risk assessment (pending improved understanding of immunity), it would be desirable to use the most specific test or that with the highest PPV, which we estimate to be SureScreen 1 band reading, followed by AbC-19. At a 20% seroprevalence, we estimate that around 1 in 20 SureScreen IgG positive readings would be a false positive. Confirmatory testing, possibly with a second LFIA, would be an option, although requires evaluation.

## Data Availability

Data are available on reasonable request to the corresponding author.

## Funding statement

The study was commissioned by the UK Government’s Department of Health and Social Care, and was funded and implemented by Public Health England, supported by the NIHR Clinical Research Network (CRN) Portfolio. HEJ, AEA, MH and IO acknowledge support from the NIHR Health Protection Research Unit in Behavioural Science and Evaluation at the University of Bristol. DW acknowledges support from the NIHR Health Protection Research Unit in Genomics and Data Enabling at the University of Warwick. STP is supported by an NIHR Career Development Fellowship (CDF-2016-09-018). Participants in the COMPARE study were recruited with the active collaboration of NHS Blood and Transplant (NHSBT) England (www.nhsbt.nhs.uk). Funding for COMPARE was provided by NHSBT and the NIHR Blood and Transplant Research Unit (BTRU) in Donor Health and Genomics (NIHR BTRU-2014-10024). DNA extraction and genotyping were co-funded by the NIHR BTRU and the NIHR BioResource (http://bioresource.nihr.ac.uk). The academic coordinating centre for COMPARE was supported by core funding from: NIHR BTRU, UK Medical Research Council (MR/L003120/1), British Heart Foundation (RG/13/13/30194; RG/18/13/33946) and the NIHR Cambridge Biomedical Research Centre (BRC). COMPARE was also supported by Health Data Research UK, which is funded by the UK Medical Research Council, Engineering and Physical Sciences Research Council, Economic and Social Research Council, Department of Health and Social Care (England), Chief Scientist Office of the Scottish Government Health and Social Care Directorates, Health and Social Care Research and Development Division (Welsh Division), Public Health Agency (Northern Ireland), British Heart Foundation, and Wellcome. JD holds a British Heart Foundation professorship and an NIHR senior investigator award. SK is funded by a BHF Chair award (CH/12/2/29428). The views expressed are those of the authors and not necessarily those of the NHS, NIHR or Department of Health and Social Care.

## Acknowledgements

We thank the following people who supported laboratory testing, data entry, and management: Jake Hall, Maryam Razaei, Nipunadi Hettiarachchi, Sarah Nalukenge, Katy Moore, Maria Bolea, Palak Joshi, Matthew Hannah, Amisha Vibhakar, Siew Lin Ngui, Amy Gentle, Honor Gartland, Stephanie Smith, Rashara Harewood, Hamish Wilson, Shabnam Jamarani, James Bull, Martha Valencia, Suzanna Barrow, Joshim Uddin, Beejal Vaghela, Shahmeen Ali. We also thank Steve Harbour and Neil Woodford, who provided staff, laboratories, and equipment; the blood donor centre staff and blood donors for participating in the COMPARE study; and Philippa Moore, Antoanela Colda and Richard Stewart for their invaluable contributions in the Milton Keynes General Hospital and Gloucestershire Hospitals study sites.

## Contributors’ statement

DW, RM, HEJ, STP, AEA, TB, AC, MH and IO planned the study. KRP and JS planned the laboratory based investigation. SK, JD, EDA, and DW planned the specificity investigations. DW, RM, EDSAB-HOME investigators and COMPARE investigators collected samples. RB, EL, and TB collated samples and performed assays. KP, JS, SN, JH, AG, HG, and SJ conducted experiments. HEJ, DW and SK did the statistical analyses. NC performed the rapid review of previous evidence. HEJ and DW wrote the paper, which all authors critically reviewed. Data have been verified by HEJ and DW.

## Declaration of Interests

JS and KP report financial activities on behalf of WHO in 2018 and 2019 in evaluation of several other rapid test kits. MH declares unrelated and unrestricted speaker fees and travel expenses in last 3 years from MSD and Gillead. JD has received grants from Merck, Novartis, Pfizer and AstraZeneca and personal fees and non-financial support from Pfizer Population Research Advisory Panel. Outside of this work, RB and EL perform meningococcal contract research on behalf of PHE for GSK, Pfizer and Sanofi Pasteur. All other authors declare no conflicts of interest.

## Notes

### Clinical Trial

ISRCTN56609224

### Clinical Protocols

http://www.isrctn.com/ISRCTN56609224

### Funding Statement

The study was commissioned by the UK Government Department of Health and Social Care, and was funded and implemented by Public Health England, supported by the NIHR Clinical Research Network (CRN) Portfolio. HEJ, AEA, MH and IO acknowledge support from the NIHR Health Protection Research Unit in Behavioural Science and Evaluation at the University of Bristol. DW acknowledges support from the NIHR Health Protection Research Unit in Genomics and Data Enabling at the University of Warwick. STP is supported by an NIHR Career Development Fellowship (CDF-2016-09-018). Participants in the COMPARE study were recruited with the active collaboration of NHS Blood and Transplant (NHSBT) England (www.nhsbt.nhs.uk). Funding for COMPARE was provided by NHSBT and the NIHR Blood and Transplant Research Unit (BTRU) in Donor Health and Genomics (NIHR BTRU-2014-10024). DNA extraction and genotyping were co-funded by the NIHR BTRU and the NIHR BioResource (http://bioresource.nihr.ac.uk). The academic coordinating centre for COMPARE was supported by core funding from: NIHR BTRU, UK Medical Research Council (MR/L003120/1), British Heart Foundation (RG/13/13/30194; RG/18/13/33946) and the NIHR Cambridge Biomedical Research Centre (BRC). COMPARE was also supported by Health Data Research UK, which is funded by the UK Medical Research Council, Engineering and Physical Sciences Research Council, Economic and Social Research Council, Department of Health and Social Care (England), Chief Scientist Office of the Scottish Government Health and Social Care Directorates, Health and Social Care Research and Development Division (Welsh Division), Public Health Agency (Northern Ireland), British Heart Foundation, and Wellcome. JD holds a British Heart Foundation professorship and an NIHR senior investigator award.  SK is funded by a BHF Chair award (CH/12/2/29428). The views expressed are those of the authors and not necessarily those of the NHS, NIHR or Department of Health and Social Care.

### Author Declarations

The study was approved by NHS Research Ethics Committee (Health Research Authority, IRAS 284980) on 02/06/2020 and PHE Research Ethics and Governance Group (REGG, NR0198) on 21/05/2020.

