## Supplementary Materials for "Accuracy of four lateral flow immunoassays for anti SARS-CoV-2 antibodies: a head-to-head comparative study"

Hayley E Jones^1*^, Ranya Mulchandani^2^, Sian Taylor-Phillips^3^, A E Ades^1^, Justin Shute^2^, Keith R Perry^2^, Nastassya L Chandra^2^, Tim Brooks^2^, Andre Charlett^2^, Matthew Hickman^1^, Isabel Oliver^2^, Stephen Kaptoge ^4^, John Danesh^4^, Emanuele Di Angelantonio^4^, COMPARE study investigators, EDSAB-HOME investigators, David Wyllie^2^

1. Population Health Sciences, Bristol Medical School, University of Bristol, UK
2. Public Health England, UK
3. University of Warwick, UK
4. University of Cambridge, UK

### Supplementary materials

#### **EDSAB-HOME further details**

EDSAB-HOME (ISRCTN56609224) was a study designed to assess the accuracy of LFIAs in key workers in England. Full details are available elsewhere (1, 2). The research protocol is available at <http://www.isrctn.com/ISRCTN56609224>

Symptom history was not part of the eligibility criteria. Recruitment was through three streams. All individuals in Streams A (fire and police officers: n = 1,147) and B (healthcare workers: n = 1,546) were recruited without regard to previous SARS-CoV-2 infection status. Stream C (n = 154) consisted of additional healthcare workers who were recruited based on self-reported previous PCR positivity. In total across the three recruitment streams, 268 individuals self-reported a previous positive PCR test. We refer to these as “known positives”. Self-reported PCR results were later validated by comparison with national laboratory records. Twelve of 268 known positives reported no symptoms. Of those reporting symptoms, the median (interquartile range) number of days between symptom onset and venesection was 63 (52 to 75).

*Sample size considerations:*

Sample size calculations are complex because of the lack of a gold standard test, and because prevalence is both unknown and increasing over time. Our approach was summarised in our research protocol (<http://www.isrctn.com/ISRCTN56609224>) as follows:

*“The following calculations assume that the laboratory-based test is 100% sensitive and 100% specific, which is known not to be the case. The calculations are therefore no more than illustrative. We assumed that the true sensitivity and specificity of the lateral flow immunoassay are both 98%. These are the minimum values currently considered acceptable by the MHRA (18/04/2020). The performance metric of the most interest is the PPV, defined as the probability that a person who tests positive does in fact have antibodies. Table S10 [of the research protocol] shows the expected 95% confidence intervals for sensitivity, specificity and PPV which would be obtained for a sample size of 1000 or 2500 participants, under various assumed values of prevalence in the study population. If we were to consider 90% PPV acceptable, and prevalence in the study sample was 20%, we would require 2,500 participants to obtain a 95% CI which was wholly above 90% PPV.*

*Test performance may vary across populations, e.g. due to variation in underlying severity of disease. To allow exploration of this, initially we proposed a cohort of 1500 healthcare workers and 1000 police officers, with later possible extension to the general public.”*

#### **Laboratory protocol**

All EDSAB-HOME samples were first analysed with two laboratory immunoassays (see main text). Choices of thresholds for dichotomisation of these two tests, and choice of Roche Elecsys as the primary immunoassay reference standard, has been described elsewhere (1, 2). Any immunoassay failing for technical reasons was repeated.

Manufacturers sent lateral flow immunoassay (LFIA) devices (Table S1) to PHE Colindale. All devices were stored in a room temperature controlled room (thresholds 16-30°C, actuals from continuous monitoring system 19-20°C). Laboratory staff reading the LFIA devices received on-site training from Abingdon Health (the manufacturers of the AbC-19 device), SureScreen and Biomerica. Laboratory staff discussed the evaluation, including use of the LFIA device, with OrientGene on a call. The manufacturers’ instructions for use were followed, with plasma being pipetted into the devices, followed by the chase buffer supplied with the kits according to the instructions for use. The first 350 COMPARE samples were interspersed randomly among EDSAB-HOME samples, with the remaining 1,650 COMPARE samples being analysed later.

Readers scored test bands using the WHO scoring system for subjectively read assay: 0 (“negative”), 1 (“very weak but definitely reactive”), 2 (“medium to strong reactivity”) or 7 (“invalid”). As this scoring system does not clearly state how to categorise “weak” bands, our readers used a score of 1 for what they considered to be either “weak” or “very weak”. The majority score across three readers for each band was taken as the consensus reading. If any band of a device was assigned a consensus score of 7 (invalid), the sample was re-tested and the re-test results taken as primary. We report numbers and proportions of invalid bands, and the total number and proportion of all devices with at least one invalid band.

We re-tested samples when LFIAs made apparent errors, as follows: for all four devices, we re-tested EDSAB-HOME samples if the result differed from an immunoassay composite reference standard of “positive on either Roche Elecsys or EuroImmun, versus negative on both”. For three of four devices, we also re-tested all COMPARE samples that tested positive, whereas for Biomerica, due to a lack of devices and a high observed false positive rate, only false positives in the first batch of 350 samples were re-tested. Any re-test results are reported as secondary.

#### **Review of previous evidence**

We searched for primary studies reporting data on the accuracy of at least one of the four lateral flow devices in detection of SARS-CoV-2 antibodies or previous SARS-CoV-2 infection. We searched Ovid MEDLINE (In-Process & Other Non-Indexed Citations and Daily), PubMed, MedRxiv/BioRxiv and Google Scholar from January 2020 to January 16 2021. Separate searches were performed for each device. Search terms included (AbC-19), (Orient Gene), (SureScreen), (Biomerica) AND ((SARS-CoV-2) OR (covid)). We also included known alternative names for these devices. Details and expanded search terms can be found in the table below. We also reviewed reference lists of relevant systematic reviews or meta-analyses.

We considered any article or pre-print published in any language. Screening was performed by a single reviewer. Studies were excluded if they did not report estimates of sensitivity or specificity for at least one of the four devices, or if they reported only meta-analysis results. Data extraction was completed by one reviewer. Information was entered into a Microsoft Excel form that collected information on the study population, sample used to evaluate the device (e.g. whole blood, plasma), specificity and sensitivity and additional free-text notes.

### Figures

#### **Figure S1:** Study flow diagram

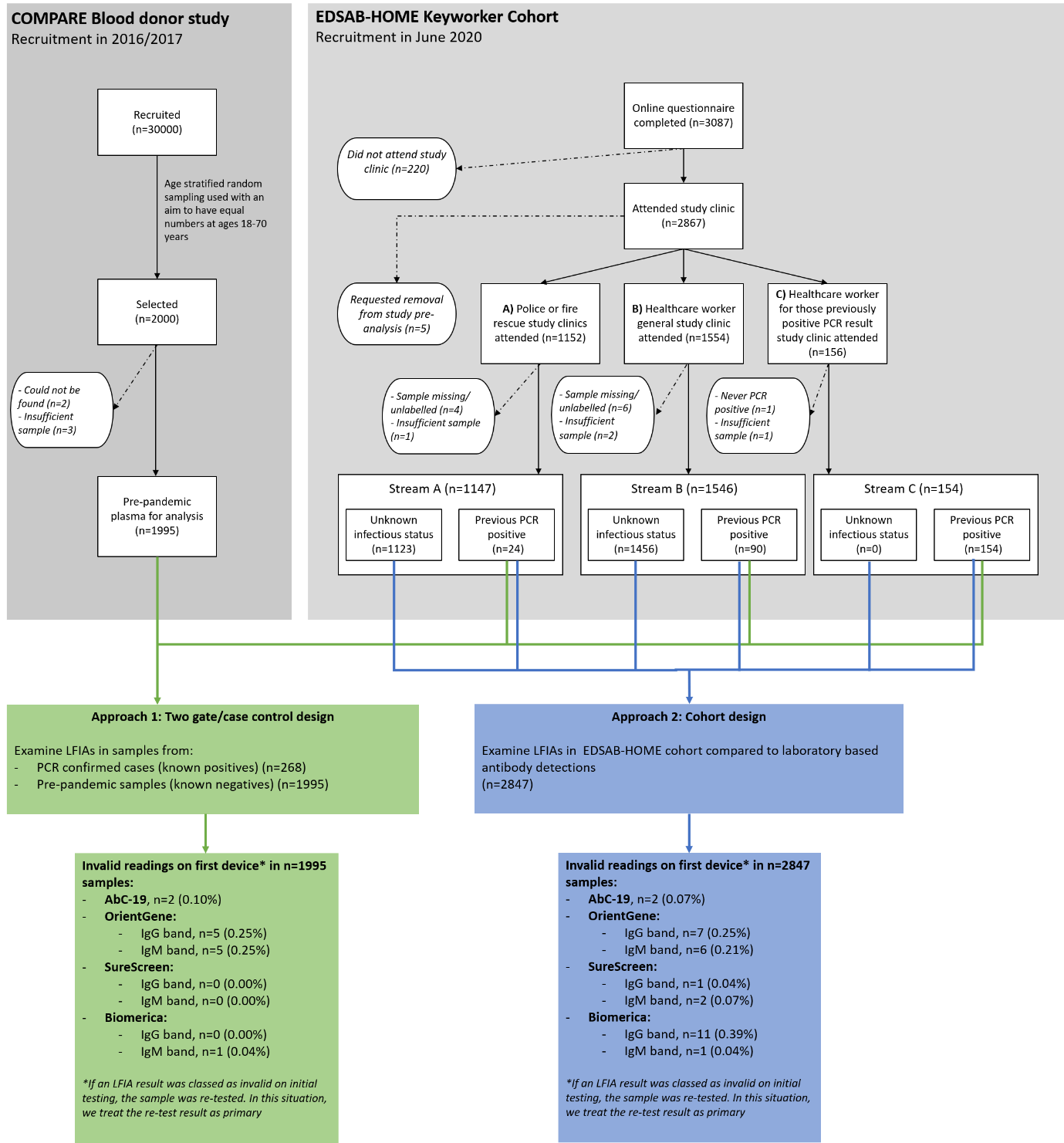

#### **Figure S2**: Relationship between specificity and age, among pre-pandemic samples

Percentage of false positive results (= 1 – specificity) by participant age at recruitment, among 1,995 pre-pandemic samples from the COMPARE study. Functional form from best fitting fractional polynomial, with adjustment for sex.

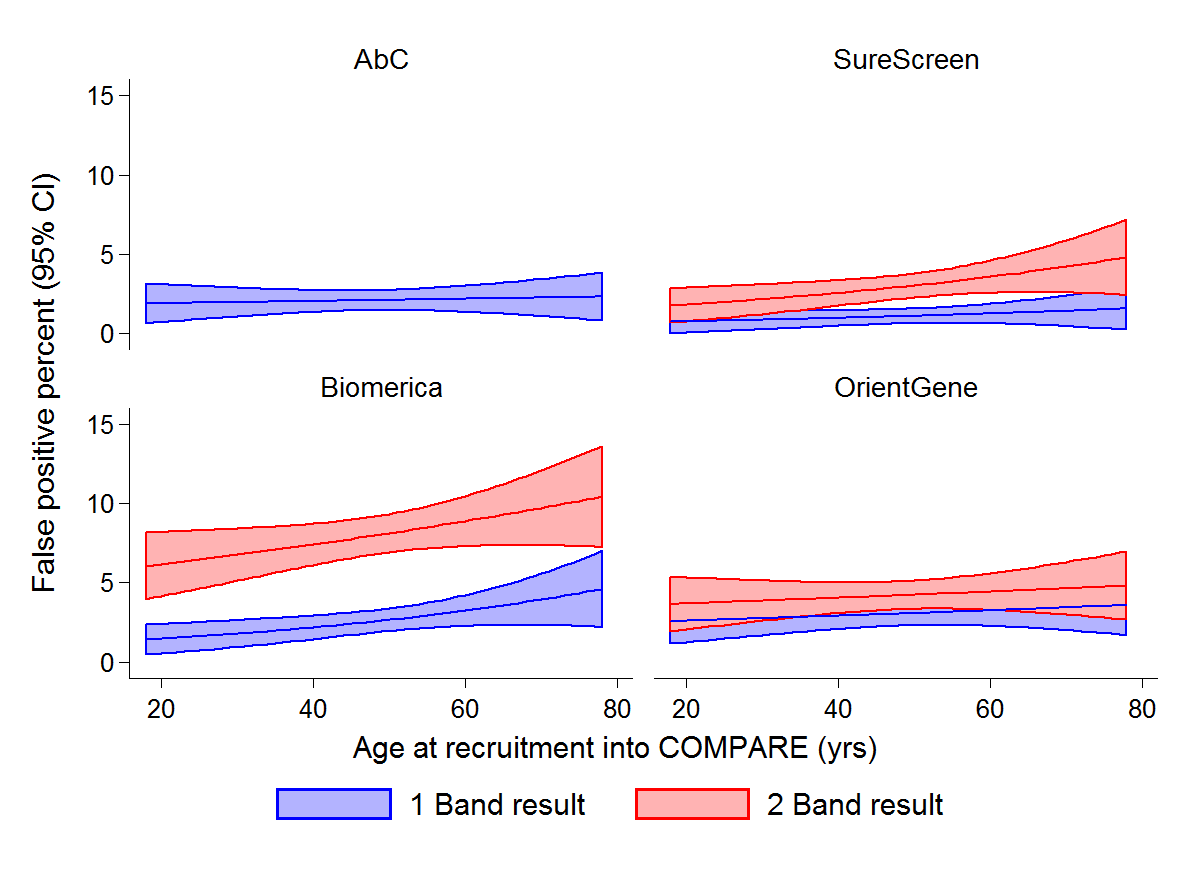

#### **Figure S3**: Lateral flow device false negativity by antibody indices

Relationship between lateral flow device detection, anti-Nucleoprotein (Roche Elecsys) and anti-Spike S1 (EuroImmun) assay results, among all 613 individuals from the EDSAB-HOME study with Roche anti-Nucleoprotein total antibody results >= 1.0 (positive).

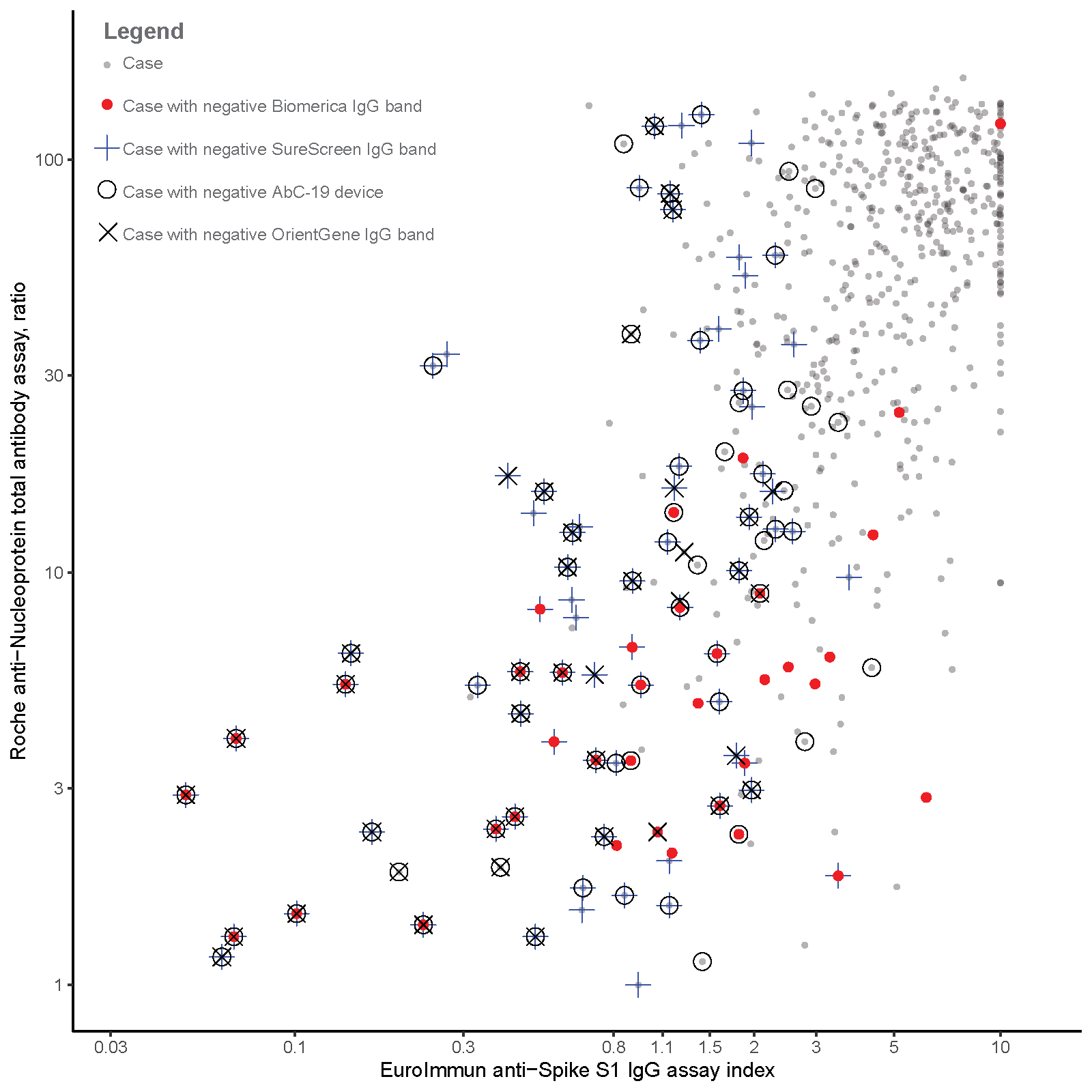

#### **Figure S4**: Positive predictive value across all pre-test probabilities

Positive predictive value (PPV) across all possible pre-test probabilities (0 to 1). PPV was estimated as a function of specificity estimated from 1,995 pre-pandemic samples (Table 1) and sensitivity from 354 Roche Elecsys positives with unknown previous infection status at clinic visit (Table 3).

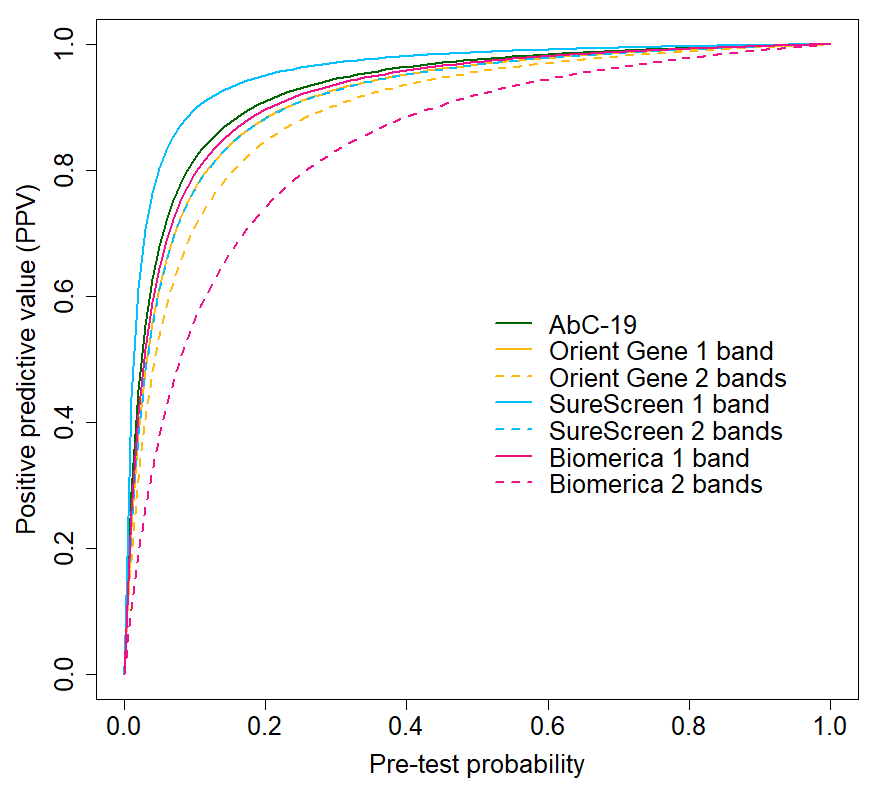

#### **Figure S5**: Review of previous evidence: workflow for literature search

Database search results, and subsequent inclusions, for rapid review of the previous evidence on the accuracy of the four lateral flow immunoassays

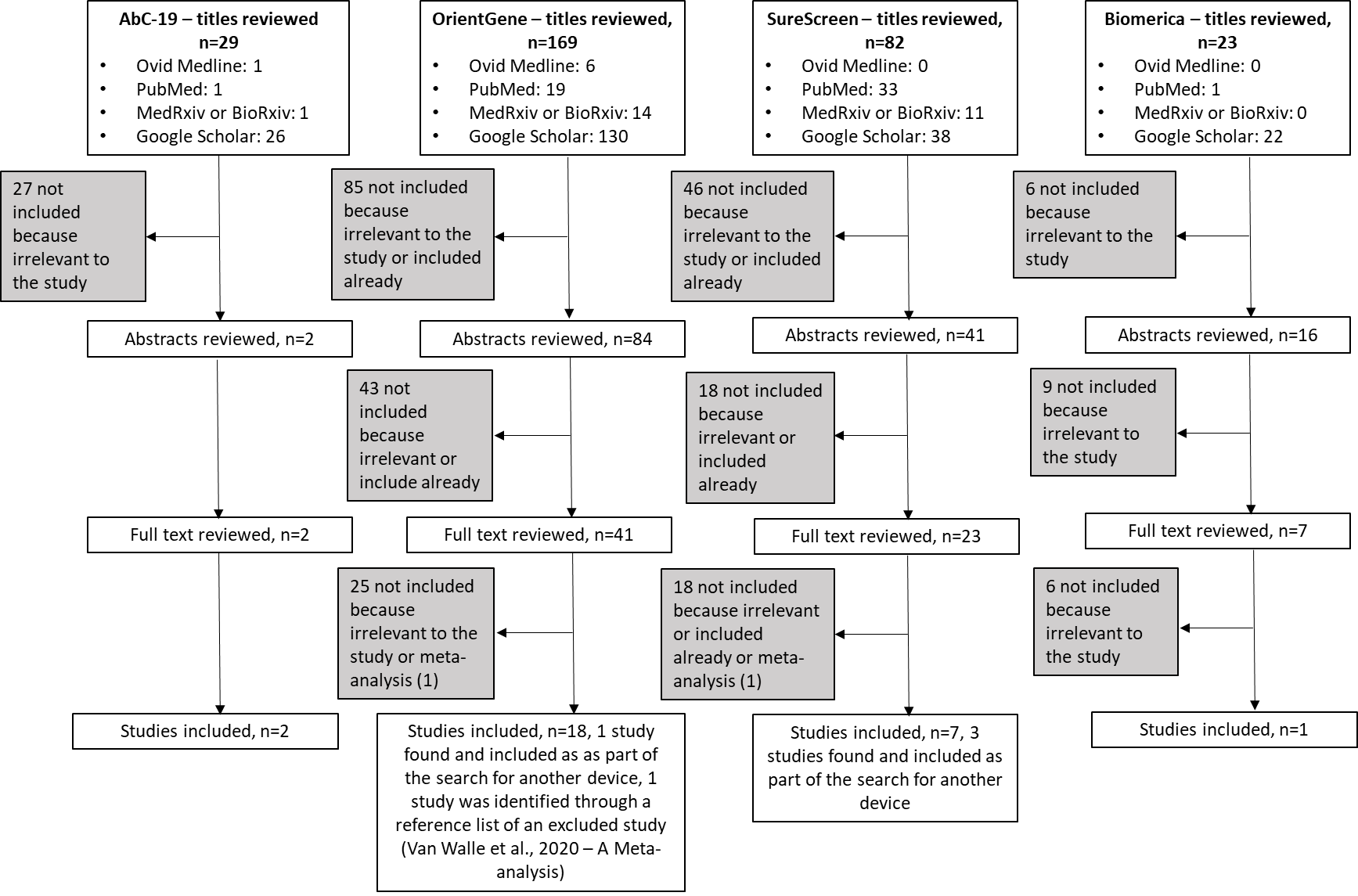

### Tables

#### **Table S1:** Lateral flow devices used

Lateral flow devices used, including the antigens contained within them and batches studied.

| Lateral flow immunoassay | Product number | Bands | Antigen used in kit | Batches evaluated | Volume of plasma added to device |
| --- | --- | --- | --- | --- | --- |
| Rapid Test Consortium “AbC-19^TM^ Rapid Test” | FG-FD51919 | IgG | Full length Spike protein (S1, S2) | A2007003 | 2.5µl |
| SureScreen “COVID-19 Rapid Test Cassette” | COVID19C | IgG+IgM | Receptor binding domain RBD (part of S1 domain of Spike protein) | COV2050005 | 10µl |
| Biomerica “COVID-19 IgG/IgM Rapid Test” | 1507A-50 | IgG+IgM | Nucleocapsid^[1]^ | 6639 | 10µl |
| Orientgene “COVID IgG/IgM Rapid Test Cassette” | GCCOV-402a | IgG+IgM | S1 domain of spike protein (Manufacturer correspondence) | 2004232  2010174  2010267  2006084 | 5µl |

#### **Table S2:** Absolute differences in specificity

Absolute differences in specificity, with 95% confidence intervals (CIs). Positive (negative) values indicate the first test has higher (lower, respectively) specificity than the second. Shaded cells: 95% CI does not include zero. CRS = composite reference standard. “Unknowns” = individuals with unknown previous infection status at clinic visit. Shaded cells: 95% CI does not include zero. CRS = composite reference standard. “Unknowns” = individuals with unknown previous infection status at clinic visit.

|  | Approach 1: known negatives (pre-pandemic) (n = 1,995) | Approach 2: “unknowns”, Roche Elecsys negative  (n = 2,225) | Approach 2: “unknowns”, EuroImmun negative (n = 2,233) | Approach 2: “unknowns”, CRS negative  (n = 2,207) |
| --- | --- | --- | --- | --- |
| AbC-19^TM^ vs Orient Gene 1 band | 0.9% (0.0, 2.0) | 1.0% (0.4, 1.7) | 1.0% (0.3, 1.7) | 0.9% (0.3, 1.6) |
| AbC-19^TM^ vs Orient Gene 2 band | 2.1% (1.0, 3.2) | 1.7% (1.0, 2.5) | 1.6% (0.9, 2.4) | 1.6% (0.9, 2.3) |
| AbC-19^TM^ vs SureScreen 1 band | -1.0% (-1.8, -0.2) | -0.1% (-0.6, 0.4) | -0.2% (-0.8, 0.3) | -0.1% (-0.7, 0.4) |
| AbC-19^TM^ vs SureScreen 2 bands | 0.9% (-0.1, 1.9) | 1.6% (0.9, 2.3) | 1.5% (0.8, 2.3) | 1.5% (0.8, 2.3) |
| AbC-19^TM^ vs Biomerica 1 band | 0.5% (-0.4, 1.5) | 1.9% (1.1, 2.7) | 2.4% (1.6, 3.2) | 2.0% (1.2, 2.9) |
| AbC-19^TM^ vs Biomerica 2 bands | 5.9% (4.6, 7.3) | 6.8% (5.7, 8.1) | 7.3% (6.1, 8.5) | 7.0% (5.9, 8.2) |
| Orient Gene 1 band vs Orient Gene 2 band | 1.1% (0.7, 1.6) | 0.7% (0.3 ,1.0) | 0.6% (0.3, 1.0) | 0.6% (0.3, 1.0) |
| Orient Gene 1 band vs SureScreen 1 band | -1.9% (-2.8, -1.1) | -1.1% (-1.8, -0.5) | -1.2% (-1.9, -0.6) | -1.1% (-1.7, -0.5) |
| Orient Gene 1 band vs SureScreen 2 bands | -0.1% (-0.9, 0.8) | 0.5% (-0.1, 1.2) | 0.5% (-0.1, 1.2) | 0.6% (-0.1, 1.3) |
| Orient Gene 1 band vs Biomerica 1 band | -0.4% (-1.5, 0.7) | 0.8% (-0.1, 1.8) | 1.4% (0.5, 2.3) | 1.1% (0.2, 2.0) |
| Orient Gene 1 band vs Biomerica 2 bands | 4.9% (3.6, 6.4) | 5.8% (4.5, 7.1) | 6.3% (5.1, 7.6) | 6.1% (4.8, 7.4) |
| Orient Gene 2 band vs SureScreen 1 band | -3.1% (-4.1, -2.2) | -1.8% (-2.5, -1.1) | -1.8% (-2.6, -1.2) | -1.7% (-2.4, -1.1) |
| Orient Gene 2 band vs SureScreen 2 bands | -1.2% (-2.2, -0.3) | -0.1% (-0.8, 0.6) | -0.1% (-0.8, 0.6) | 0.0% (-0.7, 0.6) |
| Orient Gene 2 band vs Biomerica 1 band | -1.6% (-2.7, -0.4) | 0.2% (-0.8, 1.2) | 0.8% (-0.2, 1.8) | 0.5% (-0.5, 1.4) |
| Orient Gene 2 band vs Biomerica 2 bands | 3.8% (2.3, 5.3) | 5.1% (3.8, 6.5) | 5.7% (4.4, 7.0) | 5.4% (4.1, 6.8) |
| SureScreen 1 band vs SureScreen 2bands | 1.9% (1.3, 2.6) | 1.6% (1.2, 2.2) | 1.7% (1.2, 2.3) | 1.7% (1.2, 2.3) |
| SureScreen 1 band vs Biomerica 1 band | 1.5% (0.7, 2.4) | 2.0% (1.2, 2.8) | 2.6% (1.8, 3.5) | 2.2% (1.4, 3.0) |
| SureScreen 1 band vs Biomerica 2 bands | 6.9% (5.7, 8.2) | 6.9% (5.8, 8.2) | 7.5% (6.4, 8.8) | 7.1% (6.0, 8.4) |
| SureScreen 2 band vs Biomerica 1 band | -0.4% (-1.4, 0.7) | 0.3% (-0.7, 1.3) | 0.8% (-0.1, 1.9) | 0.5% (-0.5, 1.5) |
| SureScreen 2 band vs Biomerica 2 bands | 5.0% (3.6, 6.4) | 5.2% (4.0, 6.6) | 5.8% (4.5, 7.1) | 5.5% (4.2, 6.8) |
| Biomerica 1 band vs Biomerica 2 band | 5.3% (4.4, 6.4) | 4.9% (4.1, 5.9) | 4.9% (4.0, 5.9) | 5.0% (4.1, 5.9) |

#### **Table S3:** Specificity of lateral flow devices by age

Specificity of lateral flow devices by age group, based on analysis of 1,995 “known negative” (pre-pandemic) samples. FPs = false positives. aOR = adjusted odds ratio from multivariable logistic regression with outcome of false positive and age group, sex and ethnicity (white / non-white / missing or unknown) as explanatory variables. CI = confidence interval, N/A = not applicable. *aOR per 1 year increase in age, from an alternative logistic regression with age as a continuous variable.

| **Age group** | | **<20 years** | **20-29 years** | **30-39 years** | **40-49 years** | **50-59 years** | **60-69 years** | **70+ years** | *** Linear** |
| --- | --- | --- | --- | --- | --- | --- | --- | --- | --- |
| **Number of samples** | | 27 | 295 | 365 | 383 | 362 | 412 | 151 |  |
| **AbC-19^TM^** | **FPs** | 0 | 7 | 8 | 8 | 4 | 10 | 5 |  |
|  | **Specificity** | 100% | 97.6% | 97.8% | 97.9% | 98.9% | 97.6% | 96.7% |  |
|  | **aOR**  **(95% CI)** | N/A | Ref | 0.9  (0.3,2.6) | 0.9  (0.3,2.4) | 0.5  (0.1,1.6) | 1.0  (0.4,2.7) | 1.5  (0.5,4.8) | 1.00  (0.98,1.02) |
| **Orient Gene 1 band** | **FPs** | 0 | 8 | 6 | 19 | 14 | 9 | 5 |  |
|  | **Specificity** | 100% | 97.3% | 98.4% | 95.0% | 96.1% | 97.8% | 96.7% |  |
|  | **aOR**  **(95% CI)** | N/A | Ref | 0.6  (0.2,1.8) | 1.9  (0.8,4.4) | 1.5  (0.6,3.5) | 0.8  (0.3,2.2) | 1.3  (0.4,4.0) | 1.00 (0.99,1.02) |
| **Orient Gene 2 bands** | **FPs** | 0 | 9 | 13 | 24 | 18 | 13 | 7 |  |
|  | **Specificity** | 100% | 96.9% | 96.4% | 93.7% | 95.0% | 96.8% | 95.4% |  |
|  | **aOR**  **(95% CI)** | N/A | Ref | 1.2  (0.5,2.8) | 2.2  (1.0,4.7) | 1.7  (0.7,3.8) | 1.1  (0.4,2.5) | 1.6  (0.6,4.3) | 1.00 (0.99,1.02) |
| **SureScreen 1 band** | **FPs** | 0 | 2 | 5 | 3 | 6 | 3 | 3 |  |
|  | **Specificity** | 100% | 99.3% | 98.6% | 99.2% | 98.3% | 99.3% | 98.0% |  |
|  | **aOR**  **(95% CI)** | N/A | Ref | 2.0  (0.4,10.6) | 1.2  (0.2,7.0) | 2.5  (0.5,12.4) | 1.1  (0.2,6.5) | 3.0  (0.5,18.2) | 1.01 (0.99,1.04) |
| **SureScreen 2 bands** | **FPs** | 0 | 5 | 8 | 12 | 18 | 13 | 4 |  |
|  | **Specificity** | 100% | 98.3% | 97.8% | 96.9% | 95.0% | 96.8% | 97.4% |  |
|  | **aOR**  **(95% CI)** | N/A | Ref | 1.3  (0.4,4.1) | 1.9  (0.7,5.5) | 3.0  (1.1,8.3) | 1.9  (0.7,5.5) | 1.6  (0.4,6.0) | 1.02 (1.00,1.03) |
| **Biomerica 1 band** | **FPs** | 0 | 7 | 5 | 12 | 7 | 14 | 8 |  |
|  | **Specificity** | 100% | 97.6% | 98.6% | 96.9% | 98.1% | 96.6% | 94.7% |  |
|  | **aOR**  **(95% CI)** | N/A | Ref | 0.6  (0.2,1.8) | 1.3  (0.5,3.4) | 0.8  (0.3,2.3) | 1.4  (0.6,3.5) | 2.3  (0.8,6.6) | 1.02 (1.00,1.04) |
| **Biomerica 2 bands** | **FPs** | 2 | 24 | 20 | 29 | 29 | 42 | 14 |  |
|  | **Specificity** | 92.6% | 91.9% | 94.5% | 92.4% | 92.0% | 89.8% | 90.7% |  |
|  | **aOR**  **(95% CI)** | 0.9  (0.2,4.0) | Ref | 0.7  (0.4,1.2) | 0.9  (0.5,1.6) | 1.0  (0.6,1.7) | 1.3  (0.8,2.2) | 1.2  (0.6,2.4) | 1.01 (1.00,1.02) |

#### **Table S4:** Specificity of lateral flow devices by sex and ethnicity

Specificity of lateral flow devices by sex and ethnicity based on analysis of 1,995 “known negative” (pre-pandemic) samples. FPs = false positives. aOR = adjusted odds ratio from multivariable logistic regression with outcome of false positive and age group (in 10 year bands, see Table S3), sex and ethnicity as explanatory variables. CI = confidence interval, N/A = not applicable.

| **Group** | | **Results by sex** | | **Results by ethnicity** | | |
| --- | --- | --- | --- | --- | --- | --- |
|  |  | **Male** | **Female** | **White** | **Non-white** | **Missing/unknown** |
| **Number of samples** | | 1,000 | 995 | 1,316 | 12 | 667 |
| **AbC-19^TM^** | **FPs** | 11 | 31 | 29 | 0 | 13 |
|  | **Specificity** | 98.9% | 96.9% | 97.8% | 100% | 98.1% |
|  | **aOR (95% CI)** | Ref | 2.9 (1.5, 5.9) | Ref | N/A | 1.0 (0.5, 1.9) |
| **Orient Gene 1 band** | **FPs** | 29 | 32 | 38 | 1 | 22 |
|  | **Specificity** | 97.1% | 96.8% | 97.1% | 91.7% | 96.7% |
|  | **aOR (95% CI)** | Ref | 1.1 (0.7, 1.9) | Ref | 3.0 (0.4, 24.0) | 1.2 (0.7, 2.0) |
| **Orient Gene 2 bands** | **FPs** | 40 | 44 | 53 | 1 | 30 |
|  | **Specificity** | 96.0% | 95.6% | 96.0% | 91.7% | 95.5% |
|  | **aOR (95% CI)** | Ref | 1.1 (0.7, 1.7) | Ref | 2.1 (0.3, 16.8) | 1.1 (0.7, 1.8) |
| **SureScreen 1 band** | **FPs** | 10 | 12 | 14 | 0 | 8 |
|  | **Specificity** | 99.0% | 98.8% | 98.9% | 100% | 98.8% |
|  | **aOR (95% CI)** | Ref | 1.3 (0.5, 2.9) | Ref | N/A | 1.1 (0.5, 2.7) |
| **SureScreen 2 bands** | **FPs** | 30 | 30 | 37 | 0 | 23 |
|  | **Specificity** | 97.0% | 97.0% | 97.2% | 100% | 96.6% |
|  | **aOR (95% CI)** | Ref | 1.0 (0.6, 1.7) | Ref | N/A | 1.3 (0.7, 2.1) |
| **Biomerica 1 band** | **FPs** | 21 | 32 | 38 | 0 | 15 |
|  | **Specificity** | 97.9% | 96.8% | 97.1% | 100% | 97.8% |
|  | **aOR (95% CI)** | Ref | 1.6 (0.9, 2.7) | Ref | N/A | 0.8 (0.4, 1.5) |
| **Biomerica 2 bands** | **FPs** | 73 | 87 | 102 | 1 | 57 |
|  | **Specificity** | 92.7% | 91.3% | 92.2% | 91.7% | 91.5% |
|  | **aOR (95% CI)** | Ref | 1.2 (0.9, 1.7) | Ref | 1.1 (0.1, 9.0) | 1.2 (0.8, 1.6) |

#### **Table S5:** Absolute differences in sensitivity

Absolute differences in sensitivity, with 95% confidence intervals (CIs). Positive (negative) values indicate the first test has higher (lower, respectively) sensitivity than the second. Shaded cells: 95% CI does not include zero. CRS = composite reference standard. “Unknowns” = individuals with unknown previous infection status at clinic visit. * CI was not estimated for this comparison, due to perfect correlation.

|  | Approach 1: known positives (PCR-confirmed) (n = 268) | Approach 2: “unknowns”, Roche Elecsys positive (n = 354) | Approach 2: “unknowns”, EuroImmun positive (n = 346) | Approach 2: “unknowns”, CRS positive (n = 372) |
| --- | --- | --- | --- | --- |
| AbC-19^TM^ vs Orient Gene 1 band | -1.5% (-4.4, 1.3) | -7.3% (-10.5, -4.4) | -7.7% (-11.1, -4.7) | -7.5% (-10.7, -4.5) |
| AbC-19^TM^ vs Orient Gene 2 band | -1.5% (-4.4, 1.3) | -7.6% (-10.8, -4.6) | -8.3% (-11.8, -5.2) | -8.0% (-11.3, -5.0) |
| AbC-19^TM^ vs SureScreen 1 band | 3.7% (0.5, 7.1) | -0.8% (-3.9, 2.2) | -1.7% (-4.8, 1.3) | -1.1% (-4.1, 1.9) |
| AbC-19^TM^ vs SureScreen 2 bands | -1.5% (-4.6, 1.5) | -5.0% (-8.3, -2.0) | -5.4% (-8.8, -2.3) | -5.1% (-8.2, -2.0) |
| AbC-19^TM^ vs Biomerica 1 band | -1.8% (-5.6, 1.7) | -7.0% (-10.9, -3.4) | -4.0% (-7.8, -0.4) | -5.9% (-9.7, -2.2) |
| AbC-19^TM^ vs Biomerica 2 bands | -2.6% (-6.2, 0.8) | -7.3% (-11.2, -3.6) | -4.3% (-8.1, -0.6) | -6.1% (-10.0, -2.5) |
| Orient Gene 1 band vs Orient Gene 2 band | 0.0% (N/A) * | -0.3% (-0.9, 0.3) | -0.6% (-1.4, 0.2) | -0.5% (-1.3, 0.2) |
| Orient Gene 1 band vs SureScreen 1 band | 5.1% (1.9, 8.8) | 6.4% (3.6, 9.5) | 6.0% (3.3, 9.1) | 6.4% (3.6, 9.4) |
| Orient Gene 1 band vs SureScreen 2 bands | 0.0% (-2.5, 2.5) | 2.2% (-0.1, 4.7) | 2.3% (-0.1, 4.8) | 2.4% (0.0, 4.9) |
| Orient Gene 1 band vs Biomerica 1 band | -0.4% (-3.9, 3.2) | 0.3% (-2.9, 3.5) | 3.7% (0.7, 6.9) | 1.6% (-1.6, 4.8) |
| Orient Gene 1 band vs Biomerica 2 bands | -1.1% (-4.5, 2.2) | 0.0% (-3.1, 3.1) | 3.4% (0.5, 6.6) | 1.3% (-1.8, 4.5) |
| Orient Gene 2 band vs SureScreen 1 band | 5.1% (1.9, 8.8) | 6.7% (3.9, 9.9) | 6.6% (3.7, 9.8) | 6.9% (4.0, 10.1) |
| Orient Gene 2 band vs SureScreen 2 bands | 0.0% (-2.5, 2.5) | 2.5% (0.3, 4.9) | 2.9% (0.5, 5.4) | 2.9% (0.6, 5.4) |
| Orient Gene 2 band vs Biomerica 1 band | -0.4% (-3.9, 3.2) | 0.6% (-2.6, 3.8) | 4.3% (1.2, 7.6) | 2.1% (-1.1, 5.5) |
| Orient Gene 2 band vs Biomerica 2 bands | -1.1% (-4.5, 2.2) | 0.3% (-2.9, 3.5) | 4.0% (1.0, 7.3) | 1.9% (-1.3, 5.2) |
| SureScreen 1 band vs SureScreen 2bands | -5.1% (-8.1, -2.6) | -4.2% (-6.4, -2.2) | -3.7% (-5.9, -1.8) | -4.0% (-6.1, -2.0) |
| SureScreen 1 band vs Biomerica 1 band | -5.5% (-9.6, -1.9) | -6.2% (-10.0, -2.5) | -2.3% (-6.0, 1.3) | -4.8% (-8.6, -1.1) |
| SureScreen 1 band vs Biomerica 2 bands | -6.2% (-10.3, -2.6) | -6.4% (-10.4, -2.8) | -2.6% (-6.3, 1.0) | -5.1% (-8.9, -1.4) |
| SureScreen 2 band vs Biomerica 1 band | -0.4% (-3.6, 2.8) | -2.0% (-5.5, 1.4) | 1.4% (-1.9, 4.8) | -0.8% (-4.2, 2.6) |
| SureScreen 2 band vs Biomerica 2 bands | -1.1% (-4.4, 2.0) | -2.2% (-5.7, 1.1) | 1.1% (-2.1, 4.5) | -1.1% (-4.5, 2.3) |
| Biomerica 1 band vs Biomerica 2 band | -0.7% (-1.9, 0.3) | -0.3% (-0.9, 0.3) | -0.3% (-0.9, 0.3) | -0.3% (-0.8, 0.3) |

#### **Table S6:** Sensitivity and specificity of lateral flow devices: Approach 2 with EuroImmun reference standard

Comparison with EuroImmun immunoassay in EDSAB-HOME samples, stratified by previous PCR positivity. CI = confidence interval, “Probability best” = the proportion of simulations in which the test had the highest sensitivity or specificity. Note: these AbC-19^TM^ results have been published previously (2) and are reproduced here for comparative purposes.

|  | AbC-19^TM^ | Orient Gene 1 band | Orient Gene 2 bands | SureScreen 1 band | SureScreen 2 bands | Biomerica 1 band | Biomerica 2 bands |
| --- | --- | --- | --- | --- | --- | --- | --- |
| **Analysis of 268 PCR-confirmed cases**  Reference standard of EuroImmun: 250 positive | | | | | | | |
| False negatives | 10 | 6 | 6 | 14 | 6 | 8 | 7 |
| Sensitivity (95% CI) | 96.0% (92.8, 97.8) | 97.6% (94.9, 98.9) | 97.6% (94.9, 98.9) | 94.4% (90.8, 96.6) | 97.6% (94.9, 98.9) | 96.8% (93.8, 98.4) | 97.2% (94.3, 98.6) |
| Ranked sensitivity (95% CI) | 6 (2, 7) | 2 (1, 6) | 2 (1, 6) | 7 (5, 7) | 3 (1, 6) | 5 (1, 7) | 4 (1 ,6) |
| Probability best sensitivity | 0.02 | 0.19 | 0.19 | 0.00 | 0.35 | 0.03 | 0.22 |
| **Analysis of 2,579 individuals with unknown previous infection status**  Reference standard of EuroImmun: 346 positive and 2,233 negative | | | | | | | |
| False negatives | 50 | 23 | 21 | 44 | 31 | 36 | 35 |
| Sensitivity (95% CI) | 85.5% (81.5, 88.9) | 93.4% (90.2, 95.5) | 93.9% (90.9, 96.0) | 87.3% (83.4, 90.4) | 91.0% (87.6, 93.6) | 89.6% (85.9, 92.4) | 89.9% (86.3, 92.6) |
| Ranked sensitivity (95% CI) | 7 (6, 7) | 2 (1, 3) | 1 (1, 2) | 6 (4, 7) | 3 (2, 5) | 5 (3, 6) | 4 (3, 6) |
| Probability best sensitivity | 0.00 | 0.08 | 0.91 | 0.00 | 0.01 | 0.00 | 0.00 |
| False positives | 28 | 50 | 64 | 23 | 62 | 81 | 191 |
| Specificity (95% CI) | 98.7% (98.2, 99.1) | 97.8% (97.1, 98.3) | 97.1% (96.4, 97.7) | 99.0% (98.5, 99.3) | 97.2% (96.5, 97.8) | 96.4% (95.5, 97.1) | 91.4% (90.2, 92.5) |
| Ranked specificity (95% CI) | 2 (1, 2) | 3 (3, 4) | 5 (4, 6) | 1 (1, 2) | 4 (3, 6) | 6 (5, 6) | 7 (7, 7) |
| Probability best specificity | 0.20 | 0.00 | 0.00 | 0.80 | 0.00 | 0.00 | 0.00 |

#### **Table S7:** Sensitivity and specificity of lateral flow devices: Approach 2 with composite reference standard

Sensitivity and specificity of lateral flow devices: Approach 2. Comparison with immunoassay composite reference standard (CRS) in EDSAB-HOME samples, stratified by previous PCR positivity. CI = confidence interval, “Probability best” = the proportion of simulations in which the test had the highest sensitivity or specificity. Note: these AbC-19^TM^ results have been published previously (2) and are reproduced here for comparative purposes.

|  | AbC-19^TM^ | Orient Gene 1 band | Orient Gene 2 bands | SureScreen 1 band | SureScreen 2 bands | Biomerica 1 band | Biomerica 2 bands |
| --- | --- | --- | --- | --- | --- | --- | --- |
| **Analysis of 268 PCR-confirmed cases**  Reference standard of CRS: 263 positive | | | | | | | |
| False negatives | 16 | 12 | 12 | 26 | 12 | 10 | 8 |
| Sensitivity (95% CI) | 93.9% (90.3, 96.2) | 95.4% (92.2, 97.4) | 95.4% (92.2, 97.4) | 90.1% (85.9, 93.2) | 95.4% (92.2, 97.4) | 96.2% (93.1, 97.9) | 97.0% (94.1, 98.5) |
| Ranked sensitivity (95% CI) | 6 (3, 6) | 4 (1, 6) | 4 (1, 6) | 7 (7, 7) | 3 (1, 6) | 2 (1, 6) | 1 (1, 5) |
| Probability best sensitivity | 0.01 | 0.06 | 0.06 | 0.00 | 0.10 | 0.04 | 0.73 |
| **Analysis of 2,579 individuals with unknown previous infection status**  Reference standard of CRS: 372 positive and 2,207 negative | | | | | | | |
| False negatives | 69 | 41 | 39 | 65 | 50 | 47 | 46 |
| Sensitivity (95% CI) | 81.5% (77.2, 85.1) | 89.0% (85.4, 91.8) | 89.5% (86.0, 92.2) | 82.5% (78.3, 86.0) | 86.6% (82.7, 89.7) | 87.4% (83.6, 90.4) | 87.6% (83.9, 90.6) |
| Ranked sensitivity (95% CI) | 7 (6, 7) | 2 (1, 4) | 1 (1, 3) | 6 (6, 7) | 5 (3, 5) | 4 (2, 5) | 3 (1, 5) |
| Probability best sensitivity | 0.00 | 0.06 | 0.81 | 0.00 | 0.00 | 0.02 | 0.10 |
| False positives | 21 | 42 | 56 | 18 | 55 | 66 | 176 |
| Specificity (95% CI) | 99.0% (98.5, 99.4) | 98.1% (97.4, 98.6) | 97.5% (96.7, 98.0) | 99.2% (98.7, 99.5) | 97.5% (96.8, 98.1) | 97.0% (96.2, 97.6) | 92.0% (90.8, 93.1) |
| Ranked specificity (95% CI) | 2 (1, 2) | 3 (3, 4) | 5 (4, 6) | 1 (1, 2) | 4 (3, 6) | 6 (4, 6) | 7 (7, 7) |
| Probability best specificity | 0.29 | 0.00 | 0.00 | 0.71 | 0.00 | 0.00 | 0.00 |

#### **Table S8:** Sensitivity and specificity of lateral flow devices: Approach 2 “one gate” results.

Sensitivity and specificity of lateral flow devices: Approach 2. “One gate” study results. Analysis of all EDSAB-HOME Stream A and B participants (n = 2,693). Comparison with (i) Roche Elecsys immunoassay, (ii) EuroImmun, (iii) composite immunoassay reference standard, “Probability best” = the proportion of simulations in which the test had the highest sensitivity or specificity. Note: these AbC-19^TM^ results have been published previously (2) and are reproduced here for comparative purposes.

|  | Lateral flow immunoassay | | | | | | |
| --- | --- | --- | --- | --- | --- | --- | --- |
|  | AbC-19^TM^ | Orient Gene 1 band | Orient Gene 2 bands | SureScreen 1 band | SureScreen 2 bands | Biomerica 1 band | Biomerica 2 bands |
| **Roche Elecsys reference standard:** 462 positive and 2,231 negative | | | | | | | |
| False negatives | 62 | 34 | 33 | 60 | 39 | 32 | 31 |
| Sensitivity (95% CI) | 86.6% (83.2, 89.4) | 92.6% (89.9, 94.7) | 92.9% (90.1, 94.9) | 87.0% (83.6, 89.8) | 91.6% (88.7, 93.8) | 93.1% (90.4, 95.1) | 93.3% (90.6, 95.2) |
| Ranked sensitivity (95% CI) | 7 (6, 7) | 4 (1, 5) | 3 (1, 4) | 6 (6, 7) | 5 (2, 5) | 2 (1, 5) | 1 (1, 4) |
| Probability best sensitivity | 0.00 | 0.04 | 0.32 | 0.00 | 0.02 | 0.09 | 0.53 |
| False positives | 25 | 49 | 64 | 22 | 59 | 66 | 176 |
| Specificity (95% CI) | 98.9% (98.4, 99.2) | 97.8% (97.1, 98.3) | 97.1% (96.4, 97.7) | 99.0% (98.5, 99.3) | 97.4% (96.6, 97.9) | 97.0% (96.3, 97.7) | 92.1% (90.9, 93.2) |
| Ranked specificity (95% CI) | 2 (1, 2) | 3 (3, 4) | 5 (4, 6) | 1 (1, 2) | 4 (3, 6) | 6 (3, 6) | 7 (7, 7) |
| Probability best specificity | 0.31 | 0.00 | 0.00 | 0.69 | 0.00 | 0.00 | 0.00 |
| **EuroImmun reference standard:** 451 positive and 2,242 negative | | | | | | | |
| False negatives | 57 | 27 | 25 | 50 | 35 | 40 | 39 |
| Sensitivity (95% CI) | 87.4% (84.0, 90.1) | 94.0% (91.4, 95.9) | 94.5% (91.9, 96.2) | 88.9% (85.7, 91.5) | 92.2% (89.4, 94.4) | 91.1% (88.1, 93.4) | 91.4% (88.4, 93.6) |
| Ranked sensitivity (95% CI) | 7 (6, 7) | 2 (1, 3) | 1 (1, 2) | 6 (4, 7) | 3 (2, 5) | 5 (3, 6) | 4 (3, 5) |
| Probability best sensitivity | 0.00 | 0.07 | 0.90 | 0.00 | 0.02 | 0.00 | 0.01 |
| False positives | 31 | 53 | 67 | 23 | 66 | 85 | 195 |
| Specificity (95% CI) | 98.6% (98.0, 99.0) | 97.6% (96.9, 98.2) | 97.0% (96.2, 97.6) | 99.0% (98.5, 99.3) | 97.1% (96.3, 97.7) | 96.2% (95.3, 96.9) | 91.3% (90.1, 92.4) |
| Ranked specificity (95% CI) | 2 (1, 2) | 3 (3, 4) | 5 (4, 6) | 1 (1, 2) | 4 (3, 6) | 6 (5, 6) | 7 (7, 7) |
| Probability best specificity | 0.10 | 0.00 | 0.00 | 0.90 | 0.00 | 0.00 | 0.00 |
| **Composite reference standard:** 482 positive and 2,693 negative | | | | | | | |
| False negatives | 78 | 47 | 45 | 76 | 55 | 52 | 51 |
| Sensitivity (95% CI) | 83.8% (80.3, 86.8) | 90.2% (87.3, 92.6) | 90.7% (87.7, 92.9) | 84.2% (80.7, 87.2) | 88.6% (85.4, 91.1) | 89.2% (86.1, 91.7) | 89.4% (86.4, 91.9) |
| Ranked sensitivity (95% CI) | 7 (6, 7) | 2 (1, 5) | 1 (1, 4) | 6 (6, 7) | 5 (2, 5) | 4 (1, 5) | 3 (1, 5) |
| Probability best sensitivity | 0.00 | 0.05 | 0.75 | 0.00 | 0.01 | 0.03 | 0.16 |
| False positives | 21 | 42 | 56 | 18 | 55 | 66 | 176 |
| Specificity (95% CI) | 99.1% (98.6, 99.4) | 98.1% (97.4, 98.6) | 97.5% (96.7, 98.0) | 99.2% (98.7, 99.5) | 97.5% (96.8, 98.1) | 97.0% (96.2, 97.6) | 92.0% (90.8, 93.1) |
| Ranked specificity (95% CI) | 2 (1, 2) | 3 (3, 4) | 5 (4, 6) | 1 (1, 2) | 4 (3, 6) | 6 (4, 6) | 7 (7, 7) |
| Probability best specificity | 0.29 | 0.00 | 0.00 | 0.71 | 0.00 | 0.00 | 0.00 |

#### **Table S9:** Lateral flow device results following re-tests

Proportions of re-tested samples for which the apparent error remained. (1) Reference standard of “known negative” status (pre-pandemic sample); (2) Reference standard of Roche Elecsys laboratory immunoassay. See “Laboratory protocol” for the re-testing strategy.

|  | AbC-19^TM^ | Orient Gene 1 band | Orient Gene 2 bands | SureScreen 1 band | SureScreen 2 bands | Biomerica 1 band | Biomerica 2 bands |
| --- | --- | --- | --- | --- | --- | --- | --- |
| Proportion of COMPARE (pre-pandemic) initial false positives that remained false positive on re-test (1) | 16/42  (38%) | 53/61  (87%) | 71/84  (85%) | 21/22  (96%) | 48/60  (80%) | 9/10  (90%) | 24/26  (92%) |
| Proportion of EDSAB-HOME samples that were false positive that remained false positive on re-test (2) | 8/21  (38%) | 40/42  (95%) | 54/56  (96%) | 15/18  (83%) | 52/55  (95%) | 52/66  (79%) | 114/176  (65%) |
| Proportion of EDSAB-HOME samples that were false negative that remained false negative on re-test (2) | 48/61  (79%) | 21/39  (54%) | 21/38  (55%) | 51/73  (70%) | 27/45  (60%) | 25/36  (69%) | 24/33  (73%) |

#### **Table S10:** Invalid readings

Numbers and percentages of device bands or devices with a consensus reading of “invalid”. We count a device reading as invalid overall if either band was scored as invalid. If a device was scored as invalid, the sample was re-tested and the re-tested result treated as primary.

|  | | COMPARE samples  (n = 1,995)  Number (%) | EDSAB-HOME samples  (n = 2,847)  Number (%) | Total  (n = 4,842)  Number (%) |
| --- | --- | --- | --- | --- |
| AbC-19^TM^ | | 2 (0.10%) | 2 (0.07%) | 4 (0.08%) |
| Orient Gene | IgG band | 5 (0.25%) | 7 (0.25%) | 12 (0.25%) |
|  | IgM band | 5 (0.25%) | 6 (0.21%) | 11 (0.23%) |
|  | Devices | 5 (0.25%) | 7 (0.25%) | 12 (0.25%) |
| SureScreen | IgG band | 0 (0.00%) | 1 (0.04%) | 1 (0.02%) |
|  | IgM band | 0 (0.00%) | 2 (0.07%) | 2 (0.04%) |
|  | Devices | 0 (0.00%) | 2 (0.07%) | 2 (0.04%) |
| Biomerica | IgG band | 0 (0.00%) | 11 (0.39%) | 11 (0.23%) |
|  | IgM band | 1 (0.05%) | 1 (0.04%) | 2 (0.04%) |
|  | Devices | 1 (0.05%) | 12 (0.42%) | 13 (0.27%) |

#### **Table S11:** Discordance in device band reading

Numbers and percentages (%) of qualitative disagreements on test band readings, across three expert readers in a laboratory setting. We classified readings as discordant if at least one reader scored the band as “0” (“negative”) and at least one reader scored the band as either “1” or “2”.

| Lateral flow immunoassay | AbC-19^TM^ | Orient Gene IgG band | Orient Gene IgM band | SureScreen IgG band | SureScreen IgM band | Biomerica IgG band | Biomerica IgM band |
| --- | --- | --- | --- | --- | --- | --- | --- |
| **Across all samples (n = 4,842):** | | | | | | | |
| Number | 168 | 94 | 248 | 65 | 227 | 143 | 357 |
| Percentage (95% CI) | 3.5% (3.0, 4.0) | 1.9% (1.6, 2.4) | 5.1% (4.5, 5.8) | 1.3% (1.1, 1.7) | 4.7% (4.1, 5.3) | 3.0% (2.5, 3.5) | 7.4% (6.7, 8.1) |
| **COMPARE samples (n=1,995):** | | | | | | | |
| Number | 62 | 45 | 80 | 17 | 66 | 45 | 139 |
| Percentage (95% CI) | 3.1% (2.4, 4.0) | 2.3% (1.7, 3.0) | 4.0% (3.2, 5.0) | 0.9% (0.5, 1.4) | 3.3% (2.6, 4.2) | 2.3% (1.7, 3.0) | 7.0% (5.9, 8.2) |
| **EDSAB-HOME samples that were negative on the Roche Elecsys immunoassay (n = 2,234):** | | | | | | | |
| Number | 43 | 27 | 46 | 14 | 50 | 59 | 133 |
| Percentage (95% CI) | 1.9% (1.4, 2.6) | 1.2% (0.8, 1.8) | 2.1% (1.5, 2.7) | 0.6% (0.4, 1.0) | 2.2% (1.7, 2.9) | 2.6% (2.1, 3.4) | 6.0% (5.0, 7.0) |
| **EDSAB-HOME samples that were positive on the Roche Elecsys immunoassay (n = 613):** | | | | | | | |
| Number | 63 | 22 | 122 | 34 | 111 | 39 | 85 |
| Percentage (95% CI) | 10.3% (8.1, 12.9) | 3.6% (2.4, 5.4) | 19.9% (16.9, 23.2) | 5.5% (4.0, 7.7) | 18.1% (15.3, 21.4) | 6.4% (4.7, 8.6) | 13.9% (11.4, 16.8) |

#### **Table S12:** Band strength

Consensus band scoring: “0” represents “negative”, “1” represents “very weak but definitely reactive”, “2” represents “medium to strong reactivity”.

| Band strength | Lateral flow immunoassay | | | | | | |
| --- | --- | --- | --- | --- | --- | --- | --- |
|  | AbC-19^TM^ | Orient Gene IgG band | Orient Gene IgM band | SureScreen IgG band | SureScreen IgM band | Biomerica IgG band | Biomerica IgM band |
| **Across all samples (n = 4,842):** | | | | | | | |
| 0 | 4228 (87.3%) | 4156 (85.8%) | 4360 (90.0%) | 4257 (87.9%) | 4302 (88.8%) | 4145 (85.6%) | 4489  (92.7%) |
| 1 | 343  (7.1%) | 195 (4.0%) | 366 (7.6%) | 154  (3.2%) | 366  (7.6%) | 174 (3.6%) | 321  (6.6%) |
| 2 | 271  (5.6%) | 491 (10.1%) | 116 (2.4%) | 431  (8.9%) | 174  (3.6%) | 523 (10.8%) | 32  (0.7%) |
| **COMPARE samples (n=1,995):** | | | | | | | |
| 0 | 1953 (97.9%) | 1934 (96.9%) | 1935 (97.0%) | 1973 (98.9%) | 1950 (97.7%) | 1942 (97.3%) | 1881  (94.3%) |
| 1 | 39  (2.0%) | 50  (2.5%) | 55  (2.8%) | 21  (1.1%) | 44  (2.2%) | 37  (1.9%) | 105  (5.3%) |
| 2 | 3  (0.2%) | 11  (0.6%) | 5  (0.3%) | 1  (0.1%) | 1  (0.1%) | 16  (0.8%) | 9  (0.5%) |
| **EDSAB-HOME samples that were negative on the Roche Elecsys immunoassay (n = 2,234):** | | | | | | | |
| 0 | 2206 (98.7%) | 2182 (97.7%) | 2192 (98.1%) | 2210 (98.9%) | 2188 (97.9%) | 2166 (97.0%) | 2120  (94.9%) |
| 1 | 27  (1.2%) | 39  (1.7%) | 41  (1.8%) | 17  (0.8%) | 45  (2.0%) | 46  (2.1%) | 107  (4.8%) |
| 2 | 1  (0.0%) | 13  (0.6%) | 1  (0.0%) | 7  (0.3%) | 1  (0.0%) | 22  (1.0%) | 7  (0.3%) |
| **EDSAB-HOME samples that were positive on the Roche Elecsys immunoassay (n = 613):** | | | | | | | |
| 0 | 69  (11.3%) | 40  (6.5%) | 233 (38.0%) | 74 (12.1%) | 164 (26.8%) | 37  (6.0%) | 488 (79.6%) |
| 1 | 277 (45.2%) | 106 (17.3%) | 270 (44.0%) | 116 (18.9%) | 277 (45.2%) | 91 (14.8%) | 109 (17.8%) |
| 2 | 267 (43.6%) | 467 (76.2%) | 110 (17.9%) | 423 (69.0%) | 172 (28.1%) | 485 (79.1%) | 16  (2.6%) |

#### **Table S13:** Review of previous evidence: Search terms, results and inclusions

Search strategy for rapid review of previous evidence on the accuracy of the four lateral flow immunoassays. (*) Exclusion at title stage was based on irrelevance or on previous inclusion/exclusion from an earlier search.

| Database | Search Terms | Number of titles | Number of abstracts reviewed (*) | Number of full texts reviewed | Number of papers for which data were extracted |
| --- | --- | --- | --- | --- | --- |
| Ovid Medline | ABC-19 OR UK-RTC AND SARS-CoV-2 OR covid AND limit yr of publication 2020-2021 | 1 | 1 | 1 | 1 |
| Ovid Medline | Orient Gene OR Healgen AND SARS-CoV-2 OR covid AND limit yr of publication 2020-2021 | 6 | 5 | 5 | 5 |
| Ovid Medline | Surescreen OR Sure screen OR Right Sign OR RightSign AND SARS-CoV-2 OR covid AND limit yr of publication 2020-2021 | 0 | 0 | 0 | 0 |
| Ovid Medline | Biomerica AND SARS-CoV-2 OR covid AND limit yr of publication 2020-2021 | 0 | 0 | 0 | 0 |
| PubMed | ((Abc-19) OR (UK-RTC)) AND ((SARS-CoV-2) OR (covid)) AND limit yr of publication 2020-2021 | 1 | 0 | 0 | 0 |
| PubMed | ((Orient Gene) OR (Healgen)) AND ((SARS-CoV-2) OR (covid)) AND limit yr of publication 2020-2021 | 19 | 14 | 12 | 1 |
| PubMed | ((Surescreen) OR (Sure screen) OR (Right Sign) OR (RightSign)) AND ((SARS-CoV-2) OR (covid)) AND limit yr of publication 2020-2021 | 33 | 8 | 3 | 0 |
| PubMed | (Biomerica) AND ((SARS-CoV-2) OR (covid)) AND limit yr of publication 2020-2021 | 1 | 0 | 0 | 0 |
| MedRxiv and BioRxive | "Abc-19 OR UK-RTC AND SARS-CoV-2 OR covid" and posted between "01 Jan, 2020 and 16 Jan, 2021" | 1 | 1 | 1 | 1 |
| MedRxiv and BioRxive | "Orient Gene OR OrientGene OR Healgen AND SARS-CoV-2 OR covid" and posted between "01 Jan, 2020 and 16 Jan, 2021" | 13 | 12 | 9 | 3 (NB 1 irrelevant full text was a meta-analysis: review of its reference list identified 1 additional study) |
| MedRxiv and BioRxive | “OrientGene AND SARS-CoV-2 OR covid" and posted between "01 Jan, 2020 and 16 Jan, 2021" | 1 | 1 | 1 | 0 |
| MedRxiv and BioRxive | "((Surescreen) OR (Sure screen) OR (Right Sign) OR (RightSign)) AND SARS-CoV-2 OR covid" and posted between "01 Jan, 2020 and 16 Jan, 2021" | 1 | 0 | 0 | 0 |
| MedRxiv and BioRxive | “Surescreen AND SARS-CoV-2 OR covid" and posted between "01 Jan, 2020 and 16 Jan, 2021" | 10 | 9 | 8 | 2 |
| MedRxiv and BioRxive | “Biomerica AND SARS-CoV-2 OR covid" and posted between "01 Jan, 2020 and 16 Jan, 2021" | 0 | 0 | 0 | 0 |
| Google Scholar | Abc-19 OR UK-RTC AND SARS-CoV-2 OR covid | 26 | 0 | 0 | 0 |
| Google Scholar | OrientGene AND SARS-CoV-2 OR covid | 130 | 52 | 14 | 7 |
| Google Scholar | Surescreen AND SARS-CoV-2 OR covid | 38 | 24 | 12 | 2 |
| Google Scholar | Biomerica AND SARS-CoV-2 OR covid | 22 | 16 | 7 | 1 |

#### **Table S14:** Review of previous evidence: results

Data extracted in our review of previous evidence (see “Research in Context” panel in main paper)

| Reference | Comparison with any of the other 3 devices? | Number of samples tested | Sensitivity | Specificity | Sample type | Notes |
| --- | --- | --- | --- | --- | --- | --- |
| **Rapid Test Consortium “AbC-19TM Rapid Test”** | | | | | | |
| Mulchandani (2) | No | 4,842 | See current paper | See current paper | Blood | Analysis of exact same samples as our current paper – we show these results again in the current paper, for comparison with the other devices |
| Robertson (3) | No | 818 | 97.58% (95.28, 98.95%, n = 330), estimated from samples that tested positive on EuroImmun and one other laboratory immunoassay | 99.59% (98.53, 99.95%, n = 488), estimated from pre-pandemic samples and additional samples that tested negative on 3 laboratory immunoassays | Plasma |  |
| **Orientgene “COVID IgG/IgM Rapid Test Cassette”** | | | | | | |
| Andrey (4) | No | 91 | All cases versus rIFA: whole blood = 87% (72, 95%); plasma = 97% (85, 100%)  Restricted to cases with days post diagnosis >14 days and controls (n = 77), versus rIFA = 85% (65, 95%) | All cases versus rIFA: whole blood = 98% (88, 100); plasma = 98% (88, 100%)  Restricted to cases with days post diagnosis >14 days and controls (n=77) versus rIFA = 100% (91, 100%) | Left over of whole blood EDTA samples | Population = 41 PCR-confirmed hospitalised cases + 50 negative controls (asymptomatic blood donors).  Reference standard = rIFA |
| Daoud (5) | No | 195 | IgM: 66.2% (n = 73)  IgG: 74.0% (n = 73) | IgM: 100% (n = 122)  IgG: 100% (n = 122) | Plasma from blood collected in EDTA tubes separated by centrifugation | Population = 195 patients presenting with respiratory symptoms suggestive of an infection with SARS-Cov-2. Reference standard of RT-PCR positivity. 122 were PCR negative and 73 PCR positive. |
| Herroelen (6) | No | 227 | IgM: 74.8% (67.7, 81.2%, n = 171)  IgG: 78.4% (71.4, 84.3%, n = 171) | IgM: 98.2% (90.4, 99.9, n = 56)  IgG: 94.6% (85.1, 98.9%, n = 56) | Serum | Positives are 171 sera from 135 PCR-confirmed cases |
| Dellière (7) | No | 144 | Including all samples regardless of sampling time to the onset: 93.4% (86.9, 97.3, n = 106)  Or 95.8% (89.6, 98.8) for samples collected at ≥ 10 days post symptom onset/positive PCR. | 100% (93.4, 100%, n = 42) based on pre-pandemic samples | Whole blood | Positives are 106 samples from 102 patients |
| Ong (8) | No | 278 | Overall: 43% (34, 53%, n = 99).  Sensitivity increased to 60% (46, 73%, n=52) in patients with at least 7 days of symptoms | Negatives: 98% (95, 100%, n = 129)  In the historical control sera: 100% (100, 100, n=50) | Serum | Population = patients presenting to hospital with suspected COVID-19.  228 patients and 50 sera of a historical patient control group.  NAT used as reference standard |
| Catry (9) | No | 158 | Estimates are provided stratified by patient group and days post symptom onset – see paper  Sensitivity estimated from 58 PCR confirmed cases:  Severe to critical patients (n = 18), Mild to moderate patients (n = 16), Non-hospitalised symptomatic patients (n = 24). | Combined IgM and IgG = 94 % (87.4, 97.8%), estimated from 90 pre-pandemic negative control sera, from immune or infected patients with positive Ab for various viruses (human immunodeficiency virus [HIV],  to assess the cross-reactivity. Ten samples from patients with a pathological level of rheumatoid factor (RF) (> 12 IU/mL) were included." | Serum | Positive individuals: 58 (18 severe to critical, 16 mild to moderate and 24 healthcare professionals).  Negative controls: 100 (sera from 90 with antibodies to various virus, 10 samples from high levels of rheumatoid factor) |
| Kharlamova (10) | Yes - SureScreen | 67 | N/A | 100% in IgM and IgG  Estimated from 67 pre-pandemic confounder samples with chronic inflammatory diseases | Serum |  |
| Piec (11) | No | 49 | IgG: 100% (n = 49)  Cases were 8-44 days post PCR positive test. | Not calculated (sample size too small) | Serum |  |
| Hoffman (12) | No | 300 | Lot A: IgM: 93% (85.1, 97.1%, n=74) IgG: 93.0% (86.3, 96.6%, n=100)  Lot B: IgM: 87.8% (78.5, 93.5%, n=74)  IgG: 99.5% (96.3, 100%, n=100) | Lot A: IgM: 100% (95%CI 98.1, 100%, n=200) IgG: 99.5% (97.2, 99.9%, n=200)  Lot B: IgM: 100% (98.1, 100%, n=200)  IgG: 99.5% (97.2-99.9%, n=200) | Serum | Evaluated 2 Lots of the device.  Population: Swedish COVID-19 patients or convalescents  100 positive samples confirmed by PCR (90/100) and/or serology, from Swedish COVID-19 patients or (convalescents.  200 negative controls: serum from babies (6-12 months old) and blood donor sera from 2018. |
| GeurtsvanKessel (13) | No | 99 | Overall:)  IgM: 89.4% (90.8, 95.0% n=90)  IgG: 91.5% (83.3, 96.5%, n=90).  Estimates are provided stratified by patient group and days post symptom onset – see paper | IgM = 100% (80.5, 100%, n=9)  IgG = 100% (80.5, 100%, n=9) | Serum and plasma | Specificity: Access to 147 serum and plasma, but number of sera tested for specificity was based on availability of test kit (n=9 for Orientgene)  Sensitivity: Estimated from 93 sera from 24 confirmed COVID-19 patients |
| Flower (14) | Yes – SureScreen & Biomerica | 684 | IgG band: 92.6% (86.3, 96.5%, n = 121), based on sera from PCR-confirmed cases who were also positive on composite immunoassay reference standard.  Or 89.0% (82.2, 93.8%, n = 127) based on sera from PCR-confirmed, without pre-selection. | IgG band: 97.8% (96.1, 98.9%, n = 500)  Estimated from pre-pandemic samples | Serum |  |
| Decru (15) | Yes - SureScreen | 72 | IgM in blood = 84.8%, IgM in plasma=78.8%  IgG in blood = 93.9%, IgG in plasma = 93.9%, 2 band = 97%  Estimated from 33 samples from 26 PCR confirmed cases. | IgM in blood and plasma = 100%, IgG in blood and plasma = 97.4%, 2 band in blood and plasma = 97.4%  Estimated from 39 patients without a clinical suspicion of COVID-19 who had a negative PCR result in the last 7 days | Whole blood and plasma | Positive PCR results were 21-62 days before blood sample collection. |
| Pallett (16) | No | 200 | Overall: 87% (n=150)  Subset 10-14 days post-symptom onset: 86% (73.3, 94.2%, n=50)  Subset >14 days post symptom onset: 100% (92.9, 100%, n = 50) | 96% (n=50) | Blood | Consecutive inpatients with symptoms matching PHE case definition for testing, in acute hospital.  Reference standard = RT-PCR (150 positive, 50 negative)  Estimates are provided stratified by days post symptom onset – see paper for less than 10 days |
| Alger (17) | No | 90 | Not applicable | IgM: 98.9% (94.0, 100%, n=90)  IgG: 94.4% (87.5, 98.2%, n=90) | Stored serum | Pre-pandemic (2018) serum from pregnant women |
| Van Elslande (18) | No | 138 | Estimates are provided stratified by days post symptom onset – see paper for further stratification, only presenting here day 14-25:  IgM: 97.4% (85.3, 100%, n=38)  IgG: 92.1% (78.5 98.0%, n=38);  IgM or IgG: 97.4% (85.3,100%, n=38);  IgM and IgG: 92.1% (78.5, 98.0%, n=38) | IgM: 95.1% (88.9, 98.2%, n=103)  IgG: 93.2% (86.4, 96.9%, n=103)  IgM or IgG: 91.3% (84.0,95.5%, n=103)  IgG and IgM: 97.1% (91.4, 99.4%, n=103) | Serum | Specificity: 103 samples collected before January 2020.  Sensitivity: 167 samples from 94 patients with COVID-19 confirmed with RT-PCR on nasopharyngeal swab, 38 samples used for Orientgene evaluation |
| Van Honacker (19) | No | 42 | Stratified by days post symptom onset, presented here 8-18 days:  IgM: 94% (n=23)  IgG: 88% (n=23)  IgM and IgG: 94% (n=23) | IgM: 100% (n=19)  IgG: 100% (n=19) | Serum | Sensitivity: 23 sera samples from 7 different patients admitted to hospital with PCR confirmed COVID  Estimates are provided stratified by days post symptom onset – see paper for less than 8 days  Specificity: 10 pre-pandemic sera (Aug-Sep 2019 and 9 potential cross-reactive sera (total 19)-19 |
| Pegoraro (20) | No | 149 | Stratified by days post symptom onset, presented here 14+ days:  IgG: 90% (n=21)  IgM:95% (n=21) | IgG:98% (n=67)  IgM:100% (n=67) | Serum | Population: those attending hospital emergency care or ICU with symptoms indicative of COVID-19 (Feb-Apr 2020).  Sensitivity: Estimates are provided stratified by days post symptom onset, 40 samples for 0-7 days, 21 samples for 8-13 days and 21 samples for 14+ days (total).  Specificity: 67 samples from healthy volunteers |
| Hoffman (21) | No | 153 | IgM: 69% (n=29)  IgG: 93.1% (n=29) | IgM:100% (n=124)  IgG: 99.2% (n=124) | Capillary blood samples or serum | Sensitivity: 29 PCR-confirmed COVID-19 patients or convalescents  Specificity: capillary blood samples from 124 healthy volunteers |
| **SureScreen “COVID-19 Rapid Test Cassette”** | | | | | | |
| Kharlamova (10) | Yes - OrientGene | 67 | N/A | IgM: 91% (n=67)  IgG: 91% (n=67)  Estimated from 67 pre-pandemic confounder samples with chronic inflammatory diseases | Serum |  |
| Veyrenche (22) | No | 65 | Based on 9 PCR-confirmed cases who were ≥ 14 days post symptom onset:  IgM: 78% (50.9, 100%);  IgG: 67% (36.3, 97.7%);  2 band = 78.0% (50.9, 100%) | 100% for IgM, IgG and 2 band, estimated from 20 negative controls. | Plasma | 45 PCR confirmed hospital admissions, but here we show results only for 9 who were ≥ 14 days post symptom onset. |
| Sweeney (23) | No | 601 | 94.4% (91.1-96.4%) based on 301 PCR-confirmed cases who were ≥14 days post symptom onset.  From the subset (n = 204) who were ≥ 20 days post symptom onset, sensitivity = 96.1% (92.4, 98.0%) | 99.3% (97.6, 99.8%, n = 300)  Based on 300 pre-pandemic samples: 200 stored serum samples and a panel of 100 stored acute and convalescent confounder samples | Serum |  |
| Pickering (24) | No | 160 | 80.91% (72.5, 87.16%)  Estimated from 110 samples collected from 87 patients in a hospital setting. | 100% (92.87, 100%), n = 50 | Serum | PCR-confirmed cases used to estimate sensitivity ranged from 1 to 30 days after onset of self-reported symptoms. |
| Flower (14) | Yes – OrientGene & Biomerica | 684 | IgG band: 87.5% (81.8, 91.9%, n = 184), based on sera from PCR-confirmed cases who were also positive on composite immunoassay reference standard | IgG band: 99.8% (98.9, 100%, n = 500)  Estimated from pre-pandemic samples | Serum |  |
| Decru (15) | Yes - OrientGene | 72 | IgM in blood = 87.9%, IgM in plasma = 90.9%, IgG in blood = 93.9%, IgG in plasma = 93.9%, 2 band = 97%.  Estimated from 33 samples from 26 PCR confirmed cases. | 100% for IgG, IgM and 2 band  Estimated from 39 patients without a clinical suspicion of COVID-19 who had a negative PCR result in the last 7 days. | Whole blood and plasma | Positive PCR results were 21-62 days before blood sample collection. |
| Tollånes (25) | No | 23 | IgM = 83% (70, 91%); IgG 52% (39, 65%). Estimated from n = 23 recovered outpatients with previous PCR confirmation. | N/A | Whole blood - one tube | Data extracted for study arm 2 (23 recovered outpatients with former PCR-confirmed COVID-19) only. |
| **Biomerica “COVID-19 IgG/IgM Rapid Test”** | | | | | | |
| Flower (14) | Yes – OrientGene & SureScreen | 684 | IgG band: 81.0% (74.7, 86.4%, n = 184), based on sera from PCR-confirmed cases who were also positive on composite immunoassay reference standard | IgG band: 97.8% (96.1, 98.9%, n = 500)  2 band: 96.0% (93.9, 97.5%, n = 500)  Estimated from pre-pandemic samples | Serum |  |

1. Personal communication, Freya Webber GlobalDx Ltd (distributor of the product) 20/11/2020
